# Genetic survey of biomarkers across pregnancy identifies pregnancy-specialized immune regulation

**DOI:** 10.64898/2025.12.31.25343052

**Authors:** Merve Cakir, Michela Traglia, Stacey Alexeeff, Jennifer L. Ames, Paul Ashwood, Luke P. Grosvenor, Erica P. Gunderson, Danielle HJ Kim, Jane W. Liang, Yinge Qian, Elizabeth Sahagun, Robert Yolken, Judy Van de Water, Lisa A. Croen, Lauren A. Weiss

## Abstract

Much remains unknown about the genetics of immune system changes during pregnancy. We used SNP data in a pregnancy cohort to genetically investigate 47 immune biomarkers at two timepoints, along with change between timepoints (delta). We identified 19 biomarkers with significant SNP-based heritability and 34 with genome-wide significant signals, demonstrating genetic regulation. The same biomarkers measured in early– and mid-pregnancy shared about half of significant associations across timepoints, with enrichment for immune pathways. In contrast, delta showed enrichment in transcription factors and developmental processes. About half of suggestive associations overlapped with non-pregnancy associations. However, these data leave a substantial fraction of potentially timepoint-specific and pregnancy-unique findings. Nearby genes were enriched for placental expression, reinforcing the novelty of our results. We additionally explored the relationship between immune genetic associations and prior GWAS of pregnancy complications. Overall, we present the first multi-timepoint genetic study of immune profile in pregnancy.

## INTRODUCTION

Throughout pregnancy, maternal immune cells, uterine cells, and the placenta sustain a dynamic environment to support fetal growth. The maternal immune system during pregnancy must enable implantation, prevent rejection of the semi-allogeneic fetus, and protect the mother and baby against infections^1–3^. This complex balance is maintained by fluctuations between regulatory, pro– and anti-inflammatory conditions throughout pregnancy^4,5^. During the first trimester, a pro-inflammatory environment aids implantation and placentation. As pregnancy progresses through the second trimester, the environment shifts towards immune balance and growth factors. Finally, pro-inflammatory factors become critical again in the lead up to labor. Dysregulation of this pattern can lead to pregnancy complications and hinder fetal development^3,6–8^. Therefore, insight into regulation of the maternal immune environment can inform practices that support healthy pregnancy.

Immune cells mediate the maternal environment through secretory molecules like cytokines, chemokines, and growth factors^8,9^. These factors recruit specialized immune cells, such as decidual natural killer cells and macrophages, to the maternal-fetal interface at the right time to maintain interactions between the fetus and placenta^7,10,11^. Various studies have been performed to characterize the temporal dynamics of these proteins during pregnancy. One longitudinal study revealed that abundance of 10% of 1125 plasma proteins changed significantly across gestational age^12^. These proteins were enriched in “defense response” and “leukocyte migration” pathways, highlighting immune system’s importance. Another longitudinal study that included samples obtained post-partum revealed substantial inter-individual differences that remained stable over time^13^. Despite growing knowledge of the importance of immune mediators in pregnancy, our understanding of the impact of genetics on their levels and changes during pregnancy is incomplete.

Previous studies in non-pregnant populations have identified genetic loci associated with levels of cytokines and chemokines^14,15^, highlighting the fact that genetic variation contributes to inter-individual variation and indicating potential for germline genetics to inform causal relationships. However, the immune system changes dramatically in pregnancy, suggesting that there may be genetic factors, signaling pathways, or regulatory processes exclusive to pregnancy. Our previous study investigated genetic determinants of maternal immune biomarkers at a single timepoint in the second trimester^16^. We identified three genome-wide significant (GWS) loci associated with distinct biomarkers, two of which had not been previously observed^14,16^. This work provided insight into the mid-gestational timepoint, but considering the dynamic nature of immunity, it is possible that different mechanisms would be predominant at other pregnancy stages. Therefore, studying multiple timepoints may provide valuable insight into how genetics contribute to the maternal immune milieu and differentiate between factors that impact early or mid-stages of pregnancy. Additionally, the rate at which a biomarker’s levels change could be controlled by unique regulatory mechanisms. Studies that explored response to infection or other challenges revealed different genetic factors affecting inter-individual variation at baseline and in responsiveness^17–19^. By investigating the genetic loci mediating biomarker changes, we can explore how the immune system responds to the challenge of a semi-allogeneic fetus at different phases of pregnancy.

Our previous study took advantage of genotyping and immune biomarker data in the Early Markers of Autism (EMA) study to investigate the relationship between genetics and maternal immunity^16^. Although this cohort was ascertained for children with autism spectrum disorder (ASD) and other neurodevelopmental disorders (DD), we gained novel insight into how maternal and fetal genetics shape levels of immune mediators in pregnancy. Our current IMPaCT (Immune and Metabolic Markers during Pregnancy and Child Development) study also oversamples pregnancies resulting in children with ASD and DD, and matches general population (GP) controls to these demographics^20^. Despite sampling bias, under the (testable) assumption that *genetic regulation* of the immune system is similar regardless of outcome, we can utilize this large dataset to further improve our understanding of the interplay between genetics, pregnancy, and immune system.

In this study, we identified genetic determinants of circulating levels of immune biomarkers at two different timepoints in pregnancy. To separately investigate genetic regulation of changes in biomarker levels, we also established a delta (Δ) variable based on the differences between measurements. We first established the existence of common genetic determinants using SNP-based heritability estimation and genome-wide association studies (GWAS). Next, we examined the biological processes enriched in associated loci across immune biomarkers to identify pathways important in the immune biology of early pregnancy, middle pregnancy, and responding over pregnancy. We also compared our GWAS-nominated loci with other studies of the same biomarkers in non-pregnant populations to discern putative pregnancy-specialized loci. Because the placenta is a pregnancy-exclusive organ, we examined enrichment of our loci in placental cell types to further investigate immune regulation unique to pregnancy. Finally, pregnancy complications are associated with immune system dysregulation. To investigate whether genetic drivers of the immune environment contribute to adverse pregnancy outcomes, we examined overlap between the genetics of immune biomarkers and the genetics of preeclampsia, gestational diabetes, and preterm birth.

## RESULTS

### Dataset characteristics

The IMPaCT cohort included 2436 mothers across three child outcome groups (ASD n=339, DD n=1193, GP n=904) with blood sample measurements taken at two timepoints per individual over the first and second trimesters [Fig 1a]. To study the changes observed during the progression of pregnancy, we also computed Δ based on the difference between first (T1) and second timepoint (T2) measurements.

**Fig 1.**
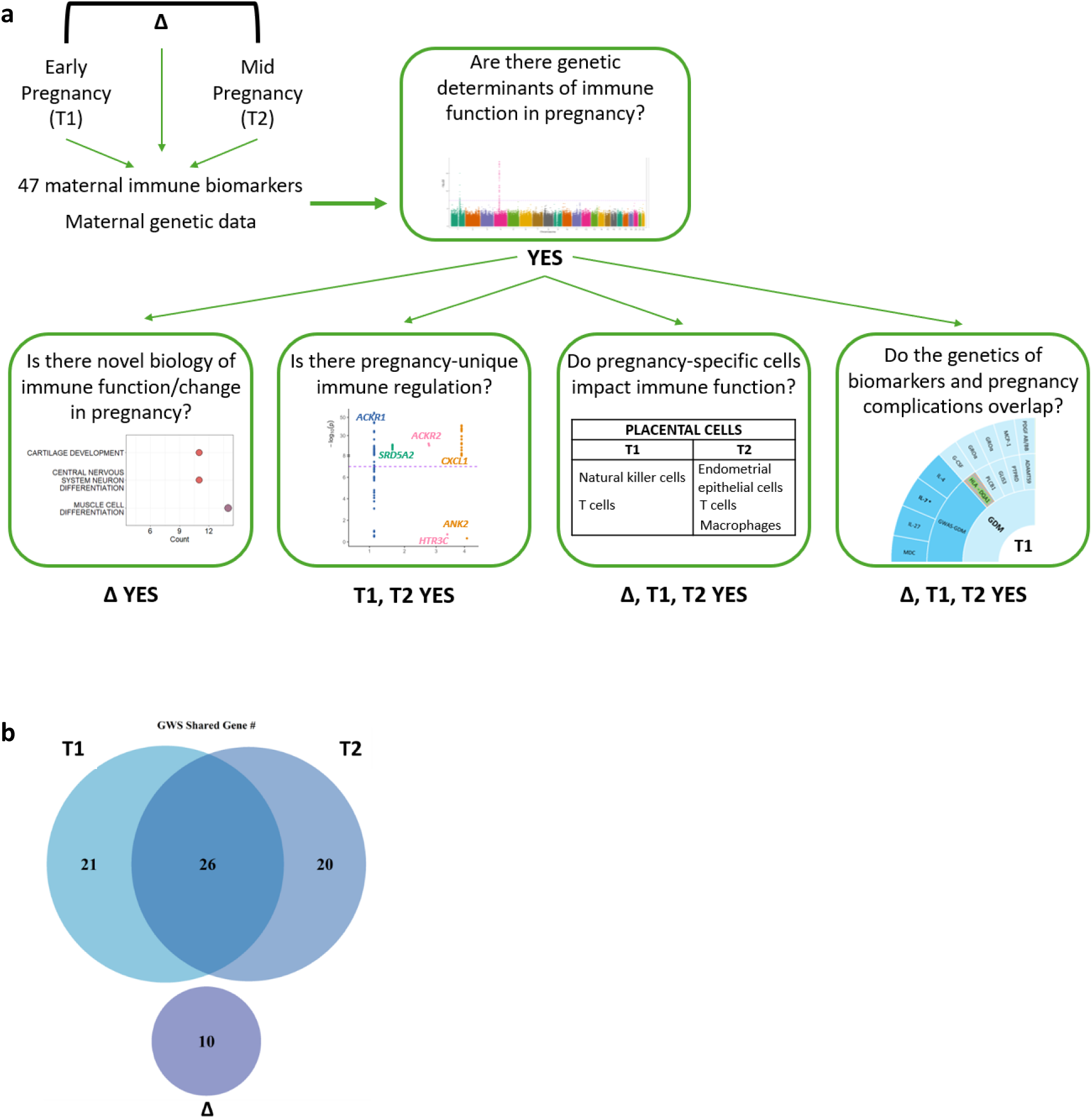
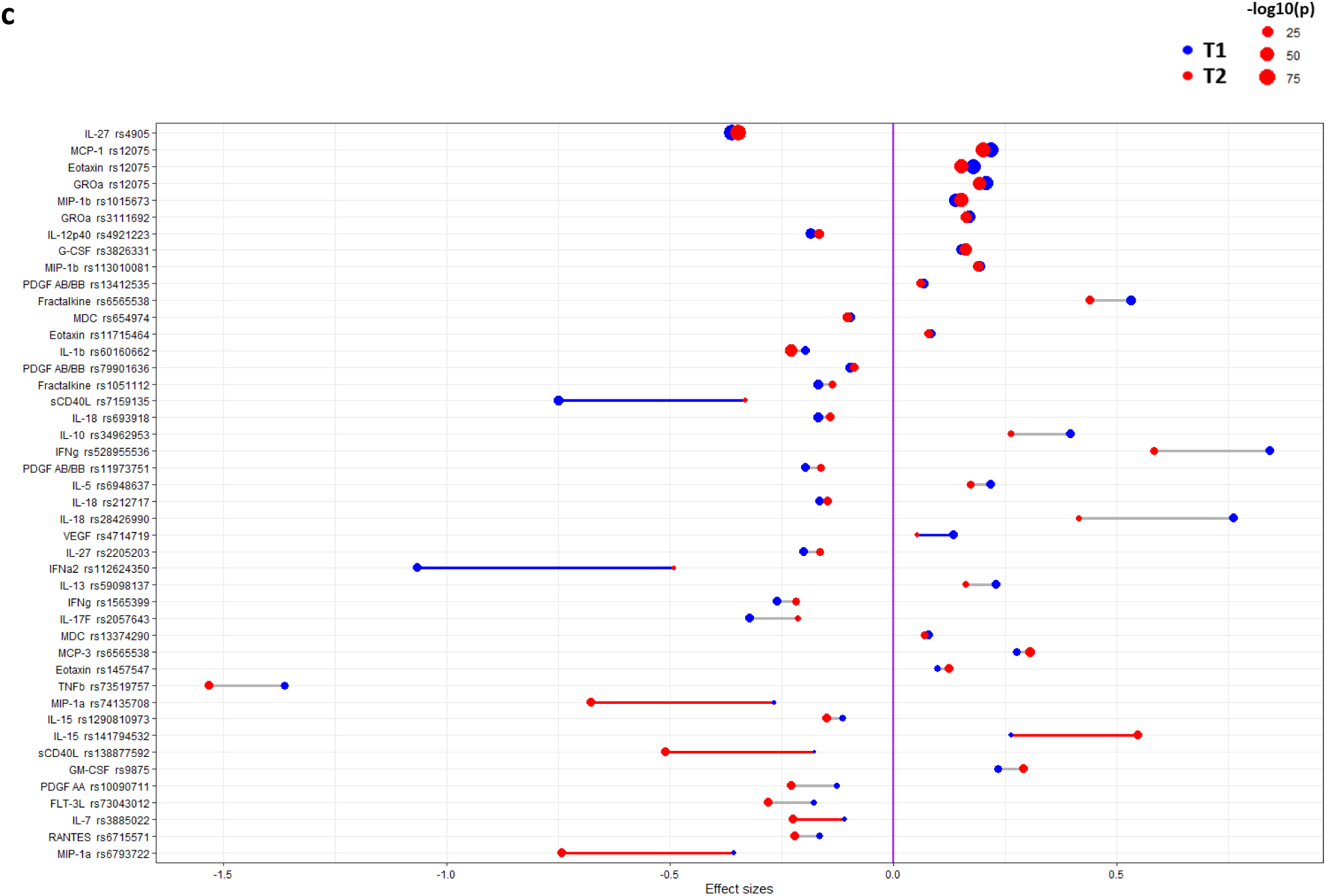
Study Design and GWAS Comparisons Across Timepoints. **a.** Schematic summarizing the study design and flow. **b.** Venn diagram of number of genes annotated to nearby GWS SNPs per time variable and overlap. **c.** T1 (blue) and T2 (red) effect size estimates of SNPs (that are GWS in at least one timepoint) are drawn with a connecting line representing the magnitude of their difference. In cases where the effect sizes are significantly different, the line is colored based on the time point with greater (absolute value) effect size, otherwise it is grey. Circle size represents the corresponding *P-value*. y axis lists the corresponding biomarker – SNP associations.

To understand the independence of our genetic results per biomarker, we computed pairwise Pearson correlation matrices per time variable [Supp Fig 1a-c]. We identified a group of 11 biomarkers that were strongly correlated across T1, T2, and Δ (FGF-2, IL-2, GM-CSF, IL-12p70, IFNγ, Fractalkine, MCP-3, IL-22, TNFβ, IL-13, TNFα). Most of the remaining biomarkers showed limited correlation: only 21 additional pairs in T1, 20 pairs in T2, and 6 pairs in Δ had r>0.5 out of 1081 pairs. Additionally, individual biomarkers’ T1-T2 measurements were highly correlated [Supp Fig 1d]. However, T1-Δ and T2-Δ for the same biomarkers showed limited correlation [Supp Fig 1e-f], suggesting that, for instance, higher T1 measurement didn’t correspond with greater decrease (or less increase) over time to regress to the mean. Likewise, inter-biomarker pairs showed some differing patterns of correlation for Δ compared with T1 or T2 [Supp Fig 1a-c]. For instance, sCD40L-EGF showed an increase in correlation in Δ (T1/T2 r=0.6, Δ r=0.84) along with a negative correlation between sCD40L-IL-7 (r=-0.35) and EGF-IL-7 (r=-0.33) not observed in T1 or T2. Overall, these phenotypic relationships suggested that genetic analyses across biomarkers and time variables could provide some independent information.

### Heritability

We estimated SNP-based heritability for all biomarkers per time variable, and in total, identified 19 biomarkers with significant heritability for at least one time variable (T1:10, T2:7, Δ:7) [Table 1]. MCP-1 had a high heritability estimate in both T1 and T2 (T1 *h^2^*=0.5, T2 *h^2^*=0.55) and PDGF AB/BB had moderate heritability in both (T1 *h^2^*=0.31, T2 *h^2^*=0.38). In contrast, there were 2 biomarkers with high (TNFβ and IL-12p70) and 5 with moderate (IL-4, fractalkine, GROα, IL-17F, IL-12p40) heritability estimates only in T1. Additionally, IL-22 had moderate estimates in both T1 and Δ (T1 *h^2^*=0.43, T2 *h^2^*=0, Δ *h^2^*=0.46), and IL-1RA (T1 *h^2^*=0.06, T2 *h^2^*=0.25, Δ *h^2^*=0.31) and IL-27 (T1 *h^2^*=0.18, T2 *h^2^*=0.3, Δ *h^2^*=0.31) had moderate estimates in T2 and Δ. IL-1α had a high heritability estimate only in Δ, along with 3 biomarkers with moderate estimates only in Δ (IL-7, IL-5, and IL-13).

**Table 1.**
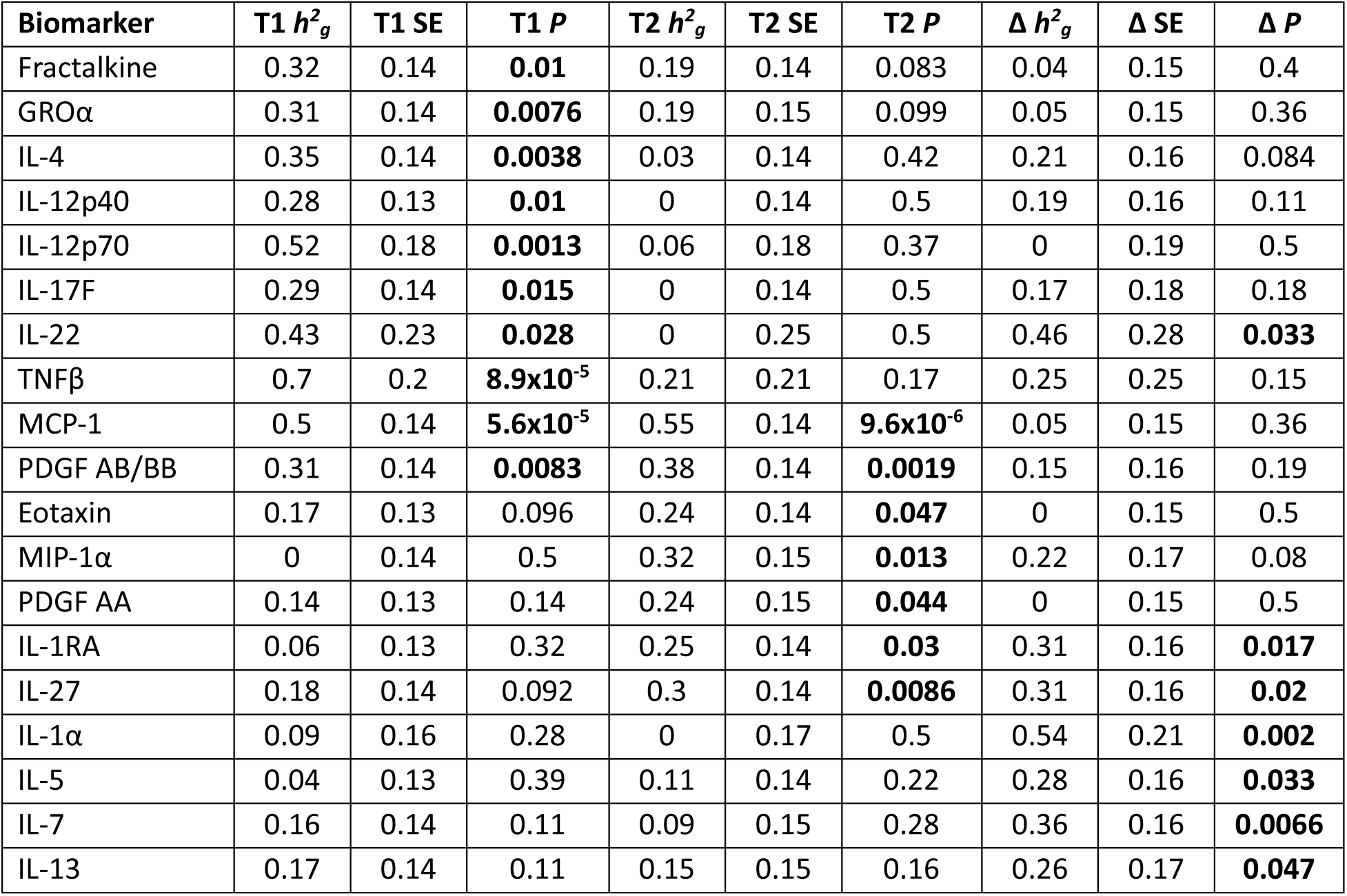
SNP-based heritability estimates per time variable. Biomarkers that have a significant estimate for at least one time variable are included. *P*<0.05 results are highlighted in bold. (*h^2^_g_*: heritability estimate, SE: standard error)

| Biomarker | T1 $h^2_g$ | T1 SE | T1 $P$ | T2 $h^2_g$ | T2 SE | T2 $P$ | $\Delta h^2_g$ | $\Delta SE$ | $\Delta P$ |
| --- | --- | --- | --- | --- | --- | --- | --- | --- | --- |
| Fractalkine | 0.32 | 0.14 | **0.01** | 0.19 | 0.14 | 0.083 | 0.04 | 0.15 | 0.4 |
| GROα | 0.31 | 0.14 | **0.0076** | 0.19 | 0.15 | 0.099 | 0.05 | 0.15 | 0.36 |
| IL-4 | 0.35 | 0.14 | **0.0038** | 0.03 | 0.14 | 0.42 | 0.21 | 0.16 | 0.084 |
| IL-12p40 | 0.28 | 0.13 | **0.01** | 0 | 0.14 | 0.5 | 0.19 | 0.16 | 0.11 |
| IL-12p70 | 0.52 | 0.18 | **0.0013** | 0.06 | 0.18 | 0.37 | 0 | 0.19 | 0.5 |
| IL-17F | 0.29 | 0.14 | **0.015** | 0 | 0.14 | 0.5 | 0.17 | 0.18 | 0.18 |
| IL-22 | 0.43 | 0.23 | **0.028** | 0 | 0.25 | 0.5 | 0.46 | 0.28 | **0.033** |
| TNFβ | 0.7 | 0.2 | **8.9x10<sup>-5</sup>** | 0.21 | 0.21 | 0.17 | 0.25 | 0.25 | 0.15 |
| MCP-1 | 0.5 | 0.14 | **5.6x10<sup>-5</sup>** | 0.55 | 0.14 | **9.6x10<sup>-6</sup>** | 0.05 | 0.15 | 0.36 |
| PDGF AB/BB | 0.31 | 0.14 | **0.0083** | 0.38 | 0.14 | **0.0019** | 0.15 | 0.16 | 0.19 |
| Eotaxin | 0.17 | 0.13 | 0.096 | 0.24 | 0.14 | **0.047** | 0 | 0.15 | 0.5 |
| MIP-1α | 0 | 0.14 | 0.5 | 0.32 | 0.15 | **0.013** | 0.22 | 0.17 | 0.08 |
| PDGF AA | 0.14 | 0.13 | 0.14 | 0.24 | 0.15 | **0.044** | 0 | 0.15 | 0.5 |
| IL-1RA | 0.06 | 0.13 | 0.32 | 0.25 | 0.14 | **0.03** | 0.31 | 0.16 | **0.017** |
| IL-27 | 0.18 | 0.14 | 0.092 | 0.3 | 0.14 | **0.0086** | 0.31 | 0.16 | **0.02** |
| IL-1α | 0.09 | 0.16 | 0.28 | 0 | 0.17 | 0.5 | 0.54 | 0.21 | **0.002** |
| IL-5 | 0.04 | 0.13 | 0.39 | 0.11 | 0.14 | 0.22 | 0.28 | 0.16 | **0.033** |
| IL-7 | 0.16 | 0.14 | 0.11 | 0.09 | 0.15 | 0.28 | 0.36 | 0.16 | **0.0066** |
| IL-13 | 0.17 | 0.14 | 0.11 | 0.15 | 0.15 | 0.16 | 0.26 | 0.17 | **0.047** |

### GWAS results

We performed GWAS for all 47 immune biomarkers at T1, T2, and Δ. 34 out of 47 biomarkers had at least one genome-wide significant (GWS) SNP (*P*<5×10^−8^) for at least one time variable: 20 biomarkers at T1 (644 SNPs), 21 at T2 (736 SNPs), and 9 for Δ (55 SNPs) [Table 2]. These GWS SNPs include highly-significant and previously-identified associations between rs4905 and IL-27 (T1 *P*=2×10^−94^, T2 *P*=4.8×10^−72^), and rs12075 and eotaxin, GROα, and MCP-1 (T1, T2 8×10^−51^>*P*>5.1×10^−20^)^14,15,21,22^. rs6565538 is associated with fractalkine and MCP-3, along with suggestive associations with GM-CSF, IFNγ, and IL-13. These 5 biomarkers are part of the 11-biomarker correlation cluster.

**Table 2.**
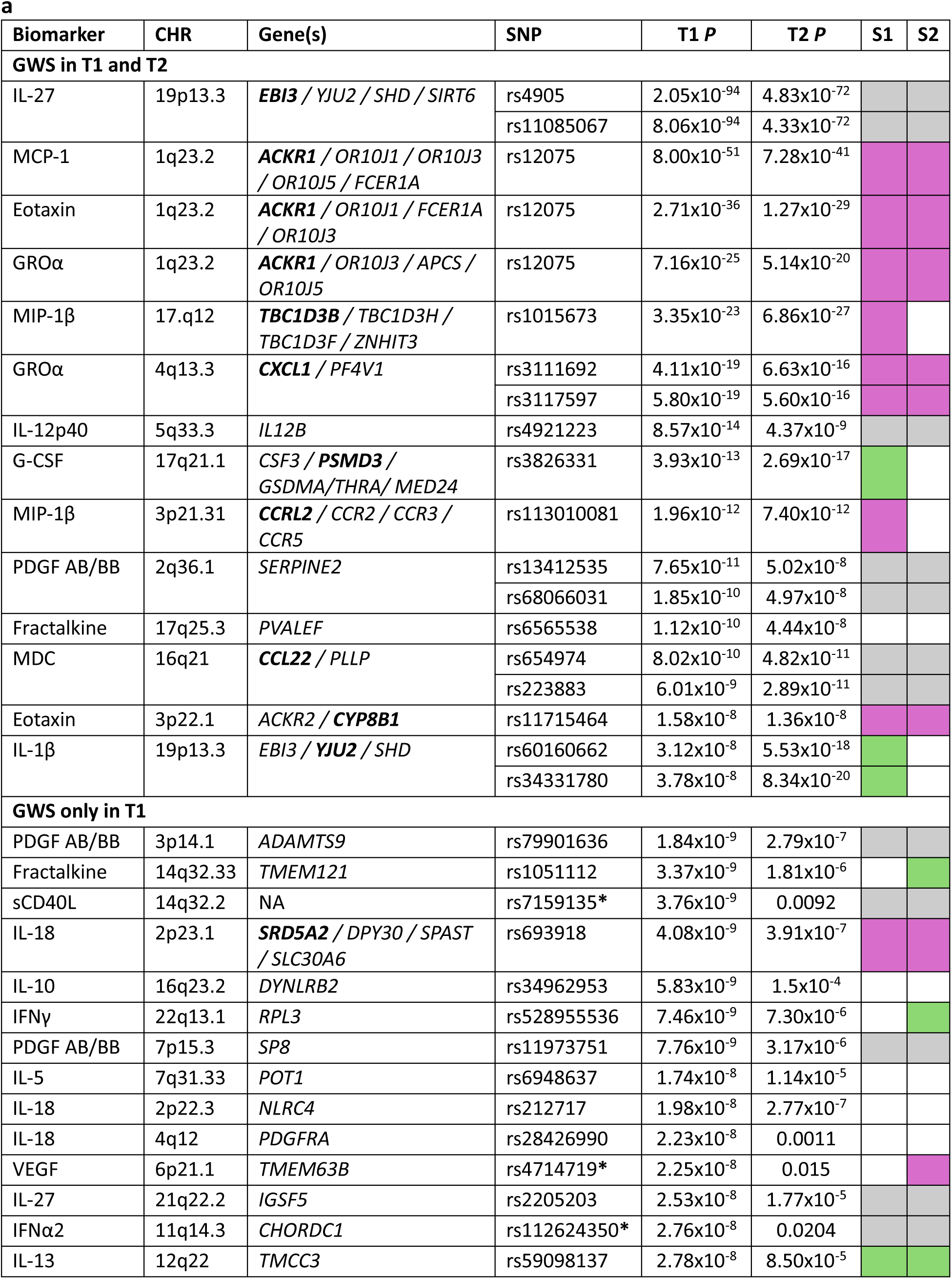
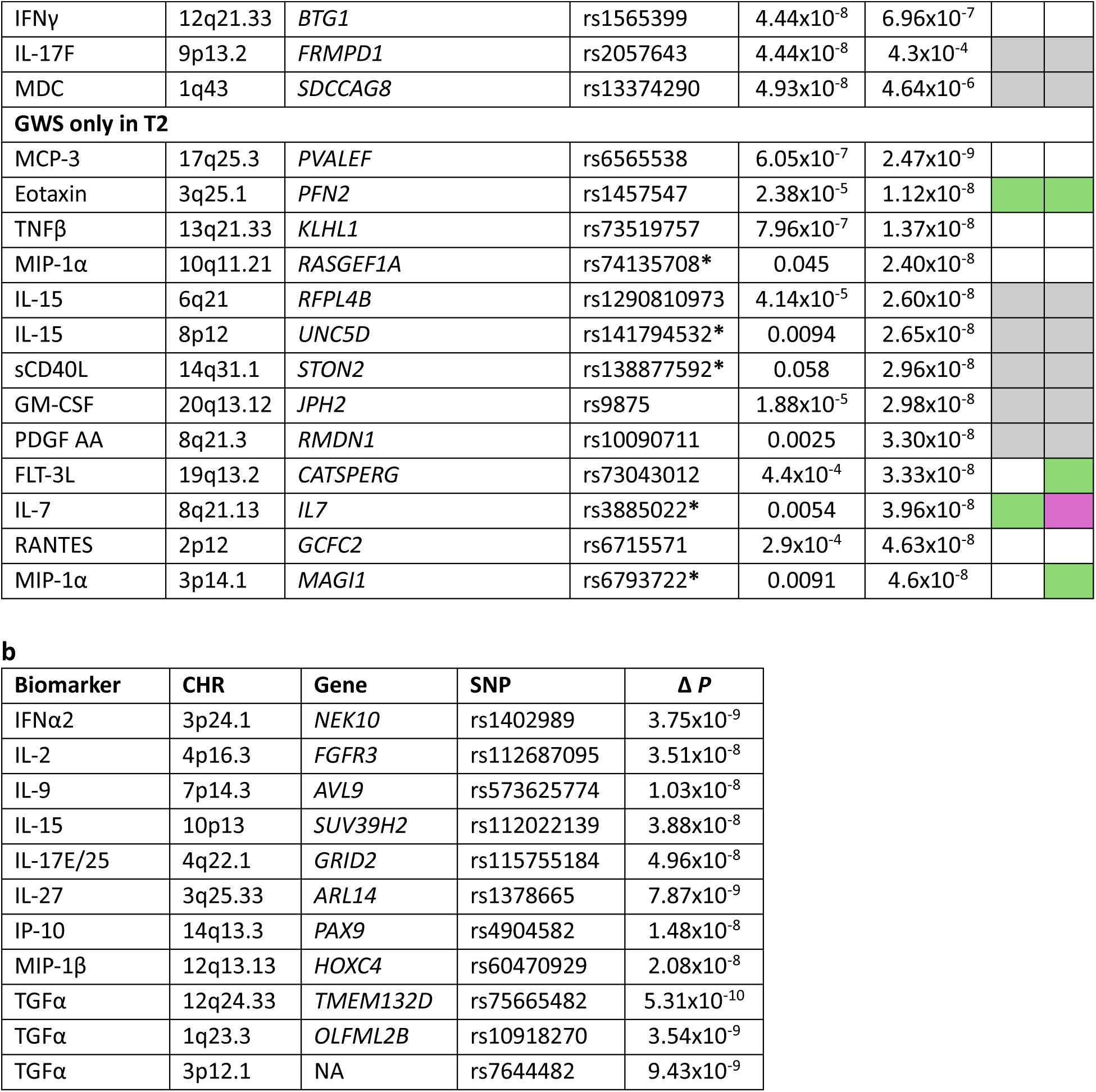
Genome-wide Significant Results. **a**. GWS loci associated with T1 and T2. If there are multiple significant SNPs within a given independent genomic region, only the most significant SNP is displayed. In cases where the most significant SNP differs between the two timepoints, both are listed in consecutive rows. Gene(s) column includes the union of all genes assigned to any GWS SNP within the genomic region, gene assigned to the SNP specified in this table is in bold. SNPs with a significant difference between T1/T2 effect size estimates are denoted with an asterisk. S1/S2 (study1/study2) column refers to the results of the comparison with non-pregnant population GWAS: pink – shared, green – unique, grey – no biomarker for comparison, white – SNP not present. **b.** GWS SNPs associated with Δ along with genes mapped to these SNPs. NA: no gene within 1Mb.

We mapped GWS SNPs to nearest genes, annotating 47 genes for T1 (1 unannotated), 46 for T2, and 10 for Δ (1 unannotated). This mapping revealed that Δ results represent a distinct set of loci, as there were no shared genes with T1 or T2 [Fig 1b]. In contrast, over half of GWS loci (26 genes) were shared between T1 and T2, the vast majority corresponding to genes that were identified as significant at both T1 and T2 for the same biomarker. These shared associations were typically autoregulatory or with their corresponding receptor, such as G-CSF–*CSF3* (rs3826331), GROα–*CXCL1* (rs3111692), MDC–*CCL22* (rs654974), and *ACKR1*’s (rs12075) associations with eotaxin, GROα, and MCP-1. However, rs4905 mapped to *EBI3* (which encodes a subunit of IL-27 and IL-35) provides an interesting example of strong association with both its own and another biomarker’s levels (IL-1β), despite modest phenotypic correlation (T1 r=0.23, T2 r=0.36). To better understand the relationship between *EBI3*, IL-27, and IL-1β, we further examined the association between the lead variant and IL-1β. After controlling for IL-27 levels, association between IL-1β and this variant was no longer significant. However, association remained highly significant with IL-27 when controlled for IL-1β.

To better characterize observed differences between T1– and T2-associated loci, we compared effect size estimates for loci GWS in at least one timepoint [Fig 1c]. In general, we observed minimal differences for loci that were GWS in both timepoints, such as IL-27–rs4905, MIP-1β–rs1015673, GROα–rs3111692, and G-CSF–rs3826331. However, associations like sCD40L–rs7159135, IL-18–rs28426990, and IFNα2–rs112624350 were GWS only in T1 and not suggestive in T2. This specificity to T1 was also reflected in the magnitude of differences between T1 and T2 effect size estimates. Similarly, there are T2-associated loci with much larger effect sizes in T2 than in T1, such as MIP-1α–rs74135708, sCD40L–rs138877592, and MIP-1α–rs6793722. Together these suggest timepoint-specific immune regulation.

Child outcome was included in our models as a covariate, as the study cohort oversampled pregnancies resulting in children with ASD and DD along with matched GP controls. To test whether the observed associations were confounded by this study design, we ran secondary analyses with GP samples only, focusing on GWS loci associated with T1, T2, and Δ. Supp Fig 2a-c shows the high correlation between effect size estimates of the full cohort results and GP-only results (T1, T2 r=0.95, Δ r=0.99), suggesting conserved genetic regulation.

**Fig 2.**
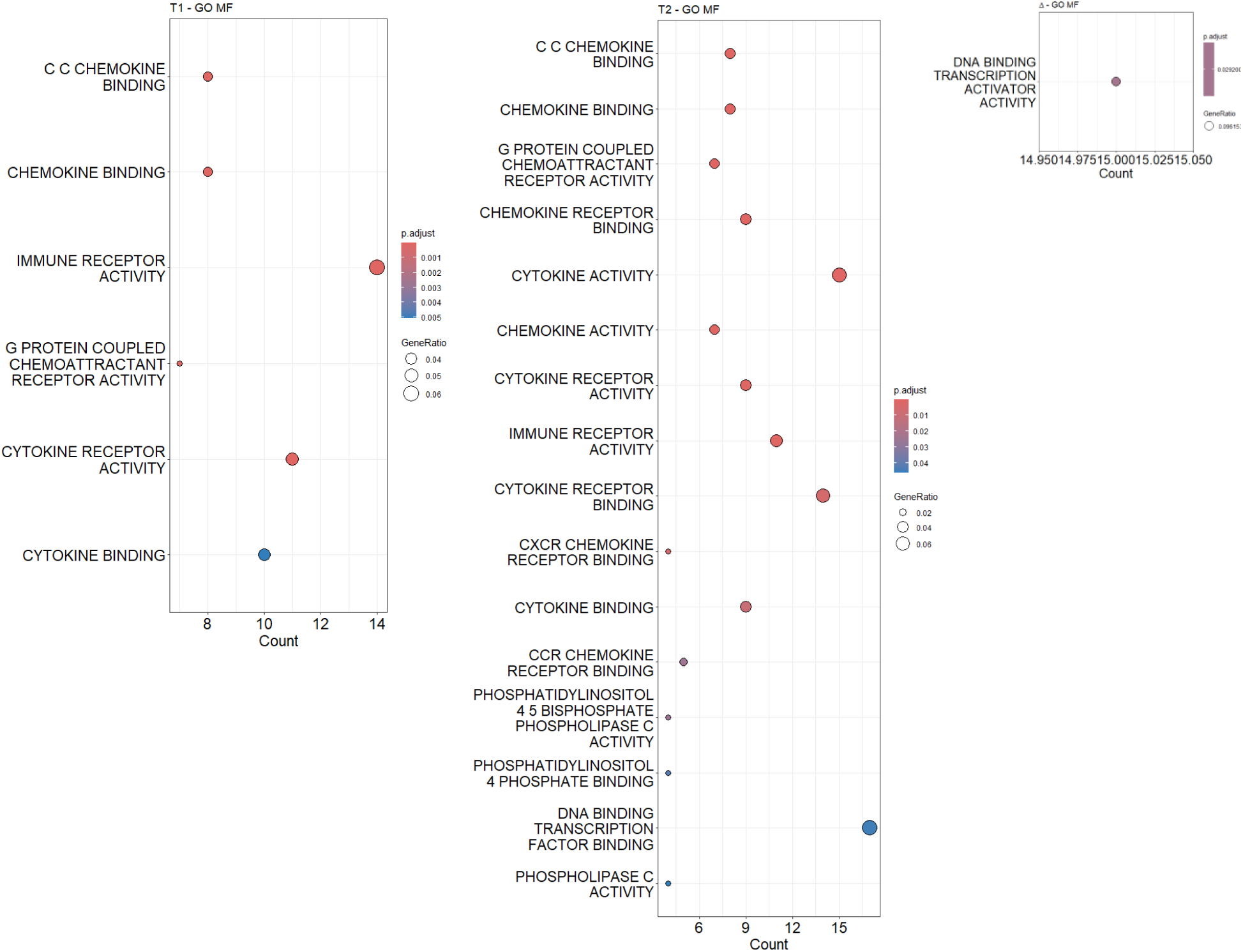
Gene Ontology Enrichment. Significantly enriched GO-MF pathways within each time variable’s set of query genes assigned to nearby suggestive SNPs (*P*<10^−6^). Categories are ordered based on adjusted p-values and circles are color coded based on adjusted p-values. Count in x axis denotes the number of genes in the query set that belong to the category. Circle sizes are proportional to gene ratio, which denotes the ratio of the number of genes in the query set in that category to the total number of genes in the category.

### Pathway enrichment analysis

To examine biological processes represented by observed associations per time variable, we performed pathway enrichment analysis after mapping suggestive SNPs (*P*<10^−6^) to nearest genes. Within each time variable, genes near suggestive SNPs across biomarkers were combined into gene sets for T1, T2, and Δ. Using GO Molecular Function (GO-MF) and GO Biological Process (GO-BP) definitions, we observed enrichment related to cytokine/chemokine binding and immune receptor activity for T1 and T2, highlighting that the associations we identified have known immune roles [Fig 2 and Supp Fig 3].

**Fig 3.**
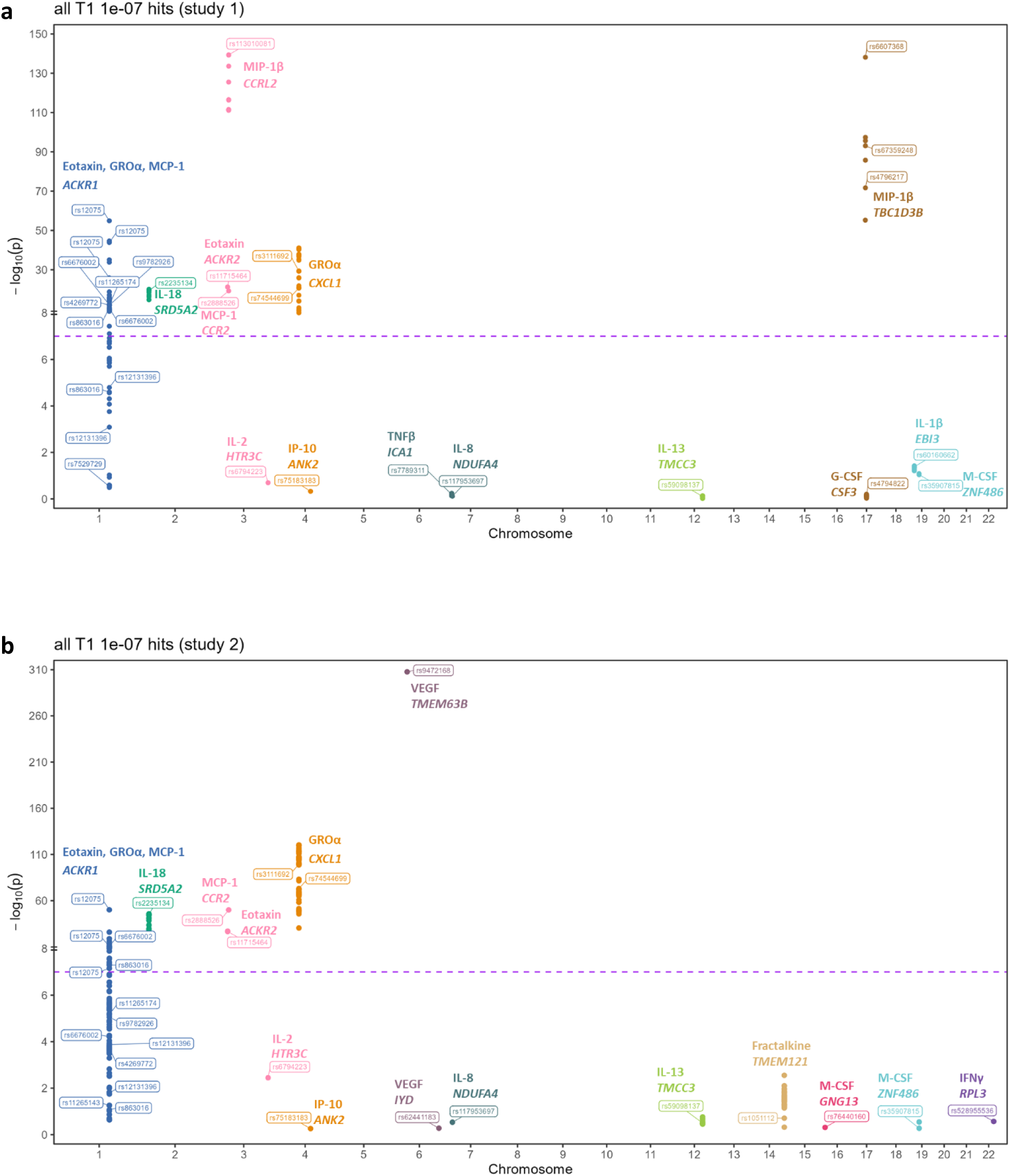
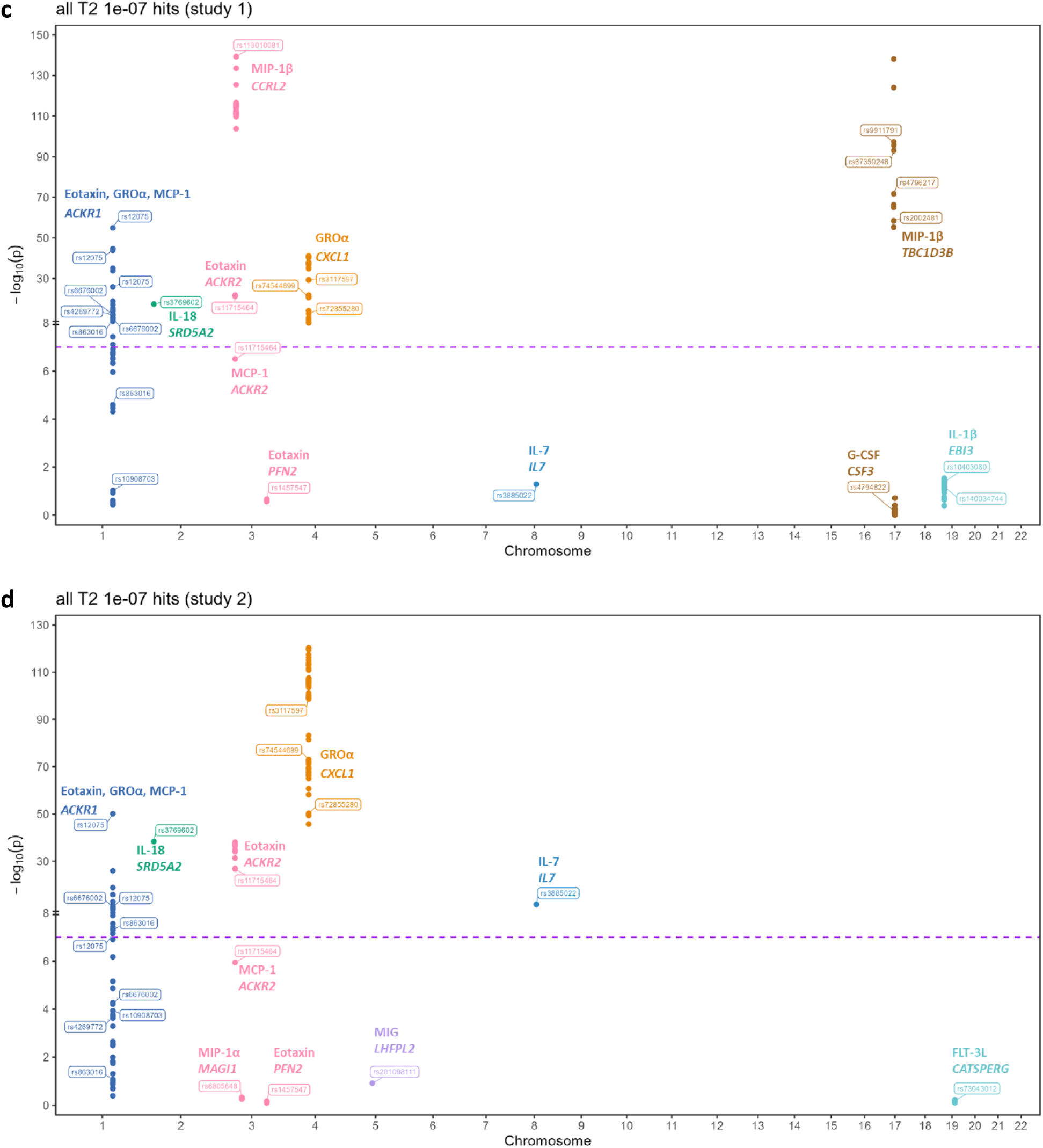
Comparison with Non-Pregnant GWAS. Highly suggestive SNPs (*P*<10^−7^) identified per biomarker are shown (dots) along with the associated biomarker and genes assigned to these variants (italicized). y axis specifies p-value in the comparison non-pregnancy study, with the purple line (10^−7^) differentiating between loci that are significant uniquely in our study or shared with the non-pregnancy study. Circles are color coded based on chromosomes. **a.** T1-associated loci compared against Study 1^14^ **b.** T1-associated loci compared against Study 2^15^ **c.** T2-associated loci compared against Study 1 **d.** T2-associated loci compared against Study 2

In line with the distinct genetic results in Δ, we identified differing enrichment. GO-BP terms show a trend towards developmental processes and differentiation [Supp Fig 3]. Transcriptional activity was the only enriched term with GO-MF [Fig 2]. Further exploration of genes associated with Δ via matched permuted results confirmed enrichment in transcription factors (*P*=0.04) [Supp Table 1]. Some of these transcription factors have known roles in embryonic development, suggesting a causal connection between variation in transcriptional developmental processes and changes in biomarker levels over pregnancy.

### Comparison with non-pregnant biomarker GWAS

Genetic regulation of immune biomarkers has previously been investigated in GWAS of non-pregnant populations^14,15^. A comparison against these studies provides the opportunity to identify variants that are particularly relevant in the context of pregnancy. For this analysis, we examined Ahola-Olli *et al.* (study 1)^14^ with 28 overlapping biomarkers and Zhao *et al.* (study 2)^15^ with 25 overlapping biomarkers. Per biomarker, we extracted highly suggestive SNPs (*P*<10^−7^) also present in the comparison study. We categorized these based on whether they are also suggestive (*P*<10^−7^) in the comparison study, labeling them as shared or unique variants, respectively. These comparisons were performed separately for study 1 and study 2.

Approximately ∼50% of T1 loci and ∼60% of T2 loci are shared with non-pregnancy studies. There is strong concordance both between the two non-pregnancy studies and also between T1 and T2 timepoints [Fig 3]; if a locus is shared between T1 and study 1, it is likely also shared between T1 and study 2 as well as with T2. For instance, the region mapped to *ACKR1* (a chemokine receptor gene) is significant in both timepoints and in both comparison studies, providing further support for the functional relationship between this receptor and eotaxin, GROα, and MCP-1 levels. One exception to this trend was observed with the VEGF–*TMEM63B* association. This region is downstream of *VEGFA* and previous studies have established that rs6921438 and additional nearby SNPs are associated with VEGF levels^23,24^. However, in our study, this association was significant only at very early stages of pregnancy, after which VEGF levels show a notable decline [Supp Fig 4], suggesting a reduced role for this variant after 50 days of gestation.

**Fig 4.**
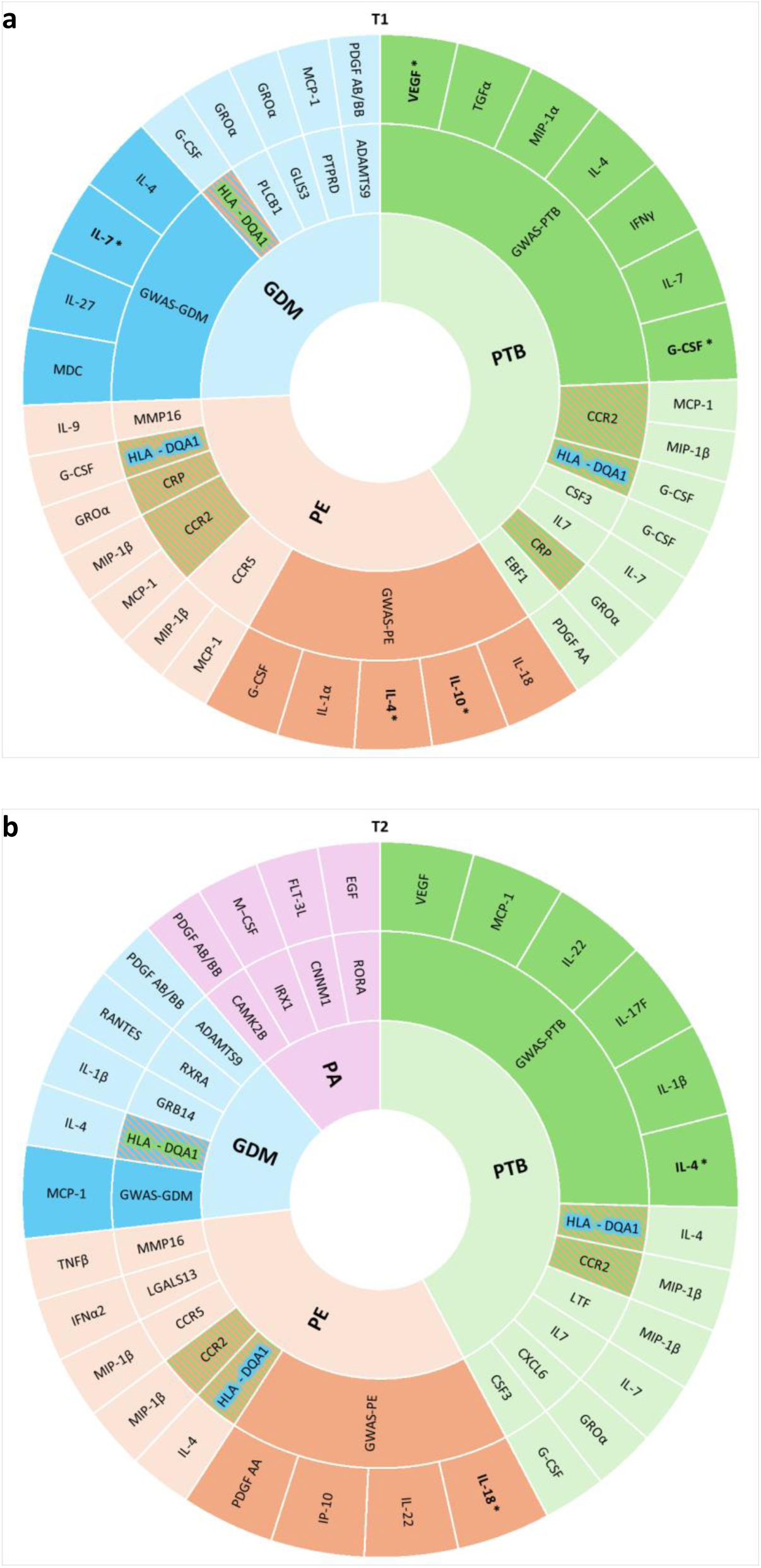
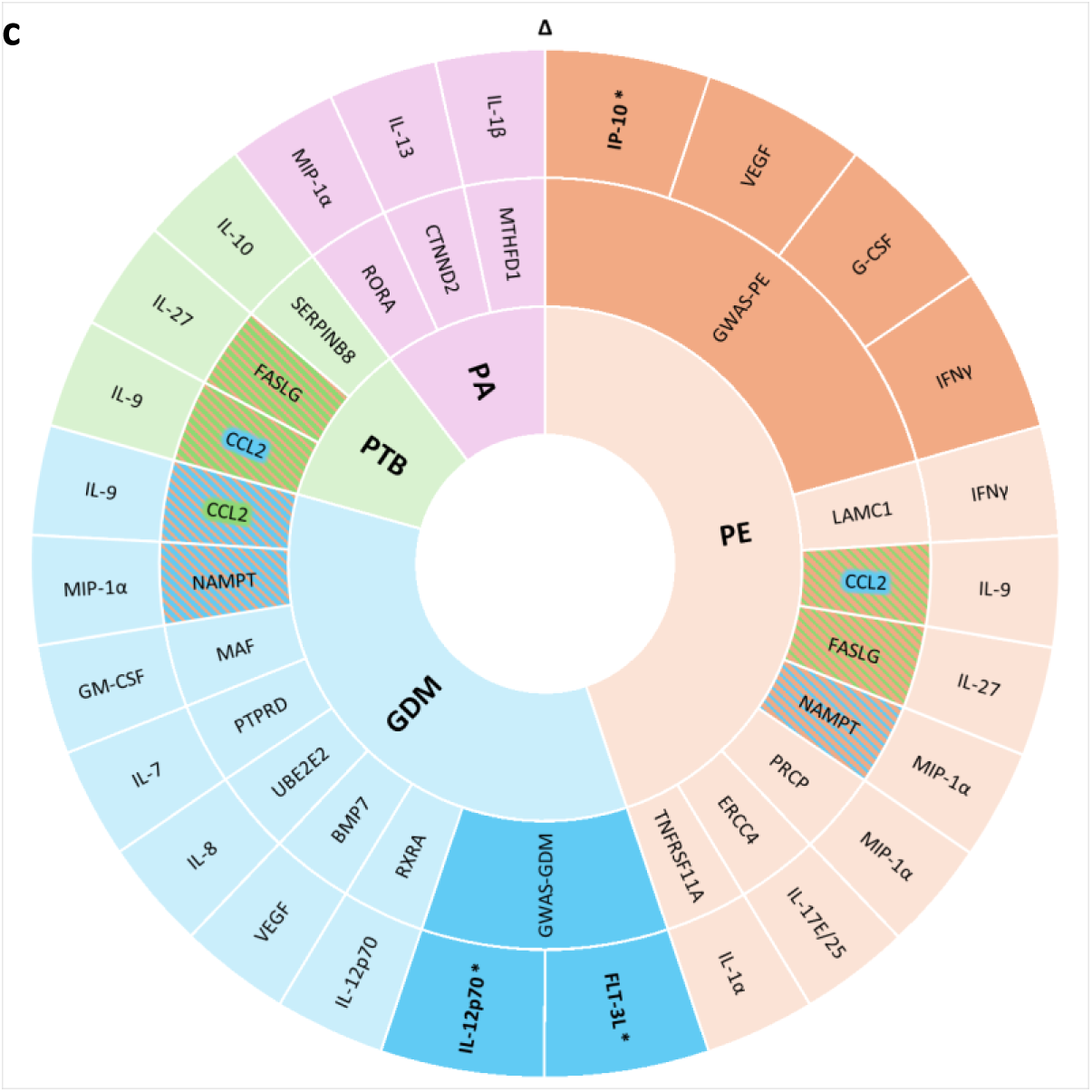
Pregnancy Complication Comparison. Four different pregnancy complications are color coded in these plots for T1 (a), T2 (b), and Δ (c): preterm birth (PTB) – green, preeclampsia (PE) – orange, gestational diabetes mellitus (GDM) – blue, and placental abruption (PA) – pink. Per condition, the outermost layer lists biomarkers of interest identified based on two separate analyses as follows. 1: The middle layer includes gene name for genes annotated to an associated SNP (*P*<10^−6^) for that time variable that were previously shown to be associated with the given condition, with the corresponding biomarker in the outer ring. Genes that are associated with multiple conditions are included in all relevant conditions and shaded with the colors of all. 2: Darker “GWAS” sections in the middle layer are attached to the biomarkers that showed significant enrichment of suggestive PE, GDM, or PTB previous GWAS SNPs (*P*<5×10^−5^) within the nominally significant SNPs (*P*<0.05) of that biomarker. (Asterisk indicates significant after multiple testing correction.) No GWAS section is included with PA as comparison against an existing study wasn’t performed and no PA layer is found in T1 as there was no overlap between PA and T1-associated genes.

Based on comparison with these two non-pregnancy studies, approximately ∼50% of our highly suggestive T1 loci and ∼40% of T2 loci are unique to our study. In cases where a given biomarker was available in both studies, loci categorized as unique based on study 1 were also consistently unique based on study 2. IL-7–*IL7* was the only exception, where it was significant in study 2 but not study 1. Unique loci include T1– and T2-associated G-CSF–*CSF3* and IL-1β–*EBI3*, as well as T1-specific (IFNγ–*RPL3*, IL-13–*TMCC3*) and T2-specific (MIP-1α–*MAGI1*, FLT-3L–*CATSPERG*, eotaxin–*PFN2*) associations.

### Placental expression enrichment

The placenta is an organ exclusive to pregnancy. Therefore, studying the gene expression patterns of placenta has the potential to determine whether our variants of interest are enriched near genes with high expression in particular placental cell types to further explore pregnancy-specialized results.

To test whether suggestive SNPs are near genes enriched in placental expression, we examined Suryawanshi *et al.*^25^, specifically expression profiles of 13 placenta-associated maternal cell types. Per cell type, we first ranked genes by expression level. Then, we compared the rank distribution of genes annotated to suggestive SNPs against genes annotated to SNPs in permuted GWAS results to determine whether placental cell types show higher than expected expression of genes near our true suggestive SNPs. Placental immune cells were relevant for T1 (natural killers) and T2 (macrophages) [Table 3]. T2 and Δ-implicated genes also showed enrichment for endometrial epithelial cells and smooth muscle cells, respectively.

**Table 3.**
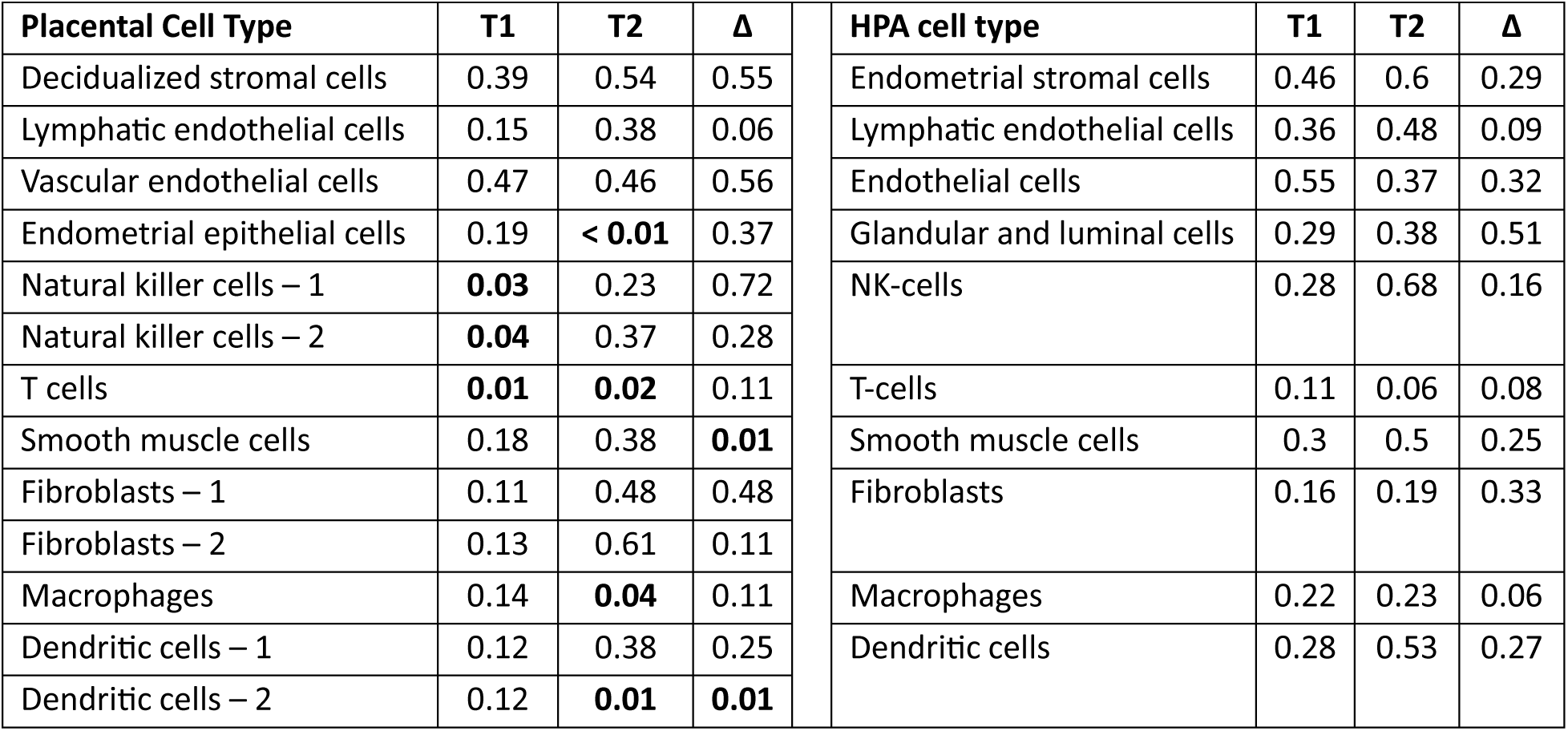
Cell Type Enrichment. Each cell type’s permutation (N=100) p-value is shown per time variable. Left half of the table lists the maternal placental cell types and the right half lists the corresponding non-placental cell types that most closely resemble their origin. *P*<0.05 results are in bold.

| Placental Cell Type | T1 | T2 | $\Delta$ | HPA cell type | T1 | T2 | $\Delta$ |
| --- | --- | --- | --- | --- | --- | --- | --- |
| Decidualized stromal cells | 0.39 | 0.54 | 0.55 | Endometrial stromal cells | 0.46 | 0.6 | 0.29 |
| Lymphatic endothelial cells | 0.15 | 0.38 | 0.06 | Lymphatic endothelial cells | 0.36 | 0.48 | 0.09 |
| Vascular endothelial cells | 0.47 | 0.46 | 0.56 | Endothelial cells | 0.55 | 0.37 | 0.32 |
| Endometrial epithelial cells | 0.19 | **< 0.01** | 0.37 | Glandular and luminal cells | 0.29 | 0.38 | 0.51 |
| Natural killer cells – 1 | **0.03** | 0.23 | 0.72 | NK-cells | 0.28 | 0.68 | 0.16 |
| Natural killer cells – 2 | **0.04** | 0.37 | 0.28 |  |  |  |  |
| T cells | **0.01** | **0.02** | 0.11 | T-cells | 0.11 | 0.06 | 0.08 |
| Smooth muscle cells | 0.18 | 0.38 | **0.01** | Smooth muscle cells | 0.3 | 0.5 | 0.25 |
| Fibroblasts – 1 | 0.11 | 0.48 | 0.48 | Fibroblasts | 0.16 | 0.19 | 0.33 |
| Fibroblasts – 2 | 0.13 | 0.61 | 0.11 |  |  |  |  |
| Macrophages | 0.14 | **0.04** | 0.11 | Macrophages | 0.22 | 0.23 | 0.06 |
| Dendritic cells – 1 | 0.12 | 0.38 | 0.25 | Dendritic cells | 0.28 | 0.53 | 0.27 |
| Dendritic cells – 2 | 0.12 | **0.01** | **0.01** |  |  |  |  |

The maternal placental cell types are highly specialized versions of blood (such as decidual natural killer cells, macrophages) and uterine (such as decidual stromal cells) cells. To test whether these cell type enrichments are specific to placenta or representing a typical expression profile of origin cells, we repeated this analysis using the single cell resource of the Human Protein Atlas^26^. The enrichments seen for placental cell types were stronger than for corresponding cell types; in all cases placental-specific enrichment was more significant in permutation testing [Table 3].

### Pregnancy complication overlaps with immune biomarker genetics

We also investigated whether the genetically-determined immune environment during pregnancy might influence the emergence of pregnancy complications. Based on literature curation in Barbitoff *et al.*, we first examined genes previously associated with the following pregnancy complications^27^: placental abruption (PA), preeclampsia (PE), gestational diabetes mellitus (GDM), and preterm birth (PTB). Across all four conditions, there were 34 genes overlapping with genes annotated to suggestive SNPs [Fig 4] (PA: 6 of 42, PE: 13 of 284, GDM: 12 of 170, PTB: 11 of 233). PA and GDM genes showed significant overlap with top immune biomarker loci (*P*=0.0042 and *P*=0.037, respectively).

For a more comprehensive and biomarker-specific view into genetic overlap between immune biomarkers and pregnancy complications, we performed comparisons against GWAS summary statistics of PE^28^, GDM^29^, and PTB^30^. We examined whether suggestive SNPs (*P*<5×10^−5^) in these studies were enriched within nominally significant SNPs (*P*<0.05) of a given biomarker in comparison to permuted results. With PE, 5 biomarkers at T1, 4 at T2, and 4 for Δ were empirically significant. IFNγ and G-CSF results are especially of interest since they are associated with PE genes (*LAMC1* and *HLA-DQA1,* respectively). The GDM comparison identified 4 biomarkers at T1, 1 at T2, and 2 for Δ. IL-12p70 was the only biomarker also associated with a known GDM gene (*RXRA*). Finally, the PTB comparison identified 7 significantly enriched biomarkers at T1 and 6 at T2. Three of these also have suggestive associations with PTB-associated genes: G-CSF (*HLA-DQA1, CSF3*), IL-7 (*IL7*), and IL-4 (*HLA-DQA1*).

## DISCUSSION

In this study, we investigated how genetics impact maternal immune biomarkers at different stages of pregnancy and within a diverse cohort of individuals. Focusing on a large set of biomarkers allowed us to ‘survey’ the maternal immune environment, identify patterns in how genetic variation acts on the immune system during pregnancy, and prioritize specific variants and loci that may have unique roles during pregnancy. Additionally, by studying the changes observed between two measurements, we identified distinct genetic determinants associated with fluctuations of biomarker levels over time.

Estimating SNP-based heritability identified 19 different heritable biomarkers, highlighting the contribution of genetic variation to the differences in immune biomarker levels observed across individuals. Interestingly, most biomarkers that showed evidence of heritability were only heritable for one time variable. This could imply that different genetic regulatory mechanisms are important (in comparison to non-genetic effects) at different timepoints and there is a dynamic interplay of genetic mechanisms over the course of pregnancy. This is surprising in light of the strong correlations between T1-T2 measurements of these biomarkers (0.78<r<0.90), and thus genetics would be acting in particular on the smaller independent portion of variation. However, we cannot disregard the possibility that limited power led to differences in detection due to noisy estimates near the threshold of significance, resulting in false negatives.

One of the strengths of our study is measurements from two timepoints in pregnancy per individual, enabling us to investigate distinct trends in early versus mid-pregnancy. Within associations categorized as unique to pregnancy, MIP-1α–*MAGI1* and eotaxin–*PFN2* were also specific to T2. These genes have previously been linked to pregnancy complications ^31,32^, suggesting that they could have specific roles during pregnancy. On the other hand, GWS SNPs shared between T1 and T2 are dominated by autoregulatory associations, such as G-CSF–*CSF3*, GROα–*CXCL1*, IL-12p40–*IL12B*, MDC–*CCL22*, and IL-27–*EBI3*. Variants near *EBI3* are especially of interest as they were also associated with IL-1β at both timepoints. The loss of association with IL-1β after controlling for IL-27 suggests that *EBI3* variants affect IL-27, which in turn affects IL-1β. Interestingly, IL-1β – *EBI3* (but not IL-27–*EBI3* ^21,22^) is also a putative pregnancy-specialized association, so the relationship between these biomarkers may change or strengthen during pregnancy.

Cross-referencing T1-vs-T2 specificity with unique-vs-shared with non-pregnancy categorization revealed a trend towards shared associations being significant in both timepoints, with VEGF being an interesting exception. The association between VEGF and rs6921438 has previously been explored^23,24^, where one study observed enhancer-associated histone marks potentially contributing to the regulation of VEGF levels^33^. The temporal trend we observed raises the possibility of this mechanism becoming reduced in saliency after ∼50 days of pregnancy. Yan *et al.* also identified this region to be associated with VEGF during pregnancy; however, they observed this association throughout first and second trimesters^34^. One key difference with our study is that their nuMoM2b cohort includes a higher percentage of pregnancies with complications, such as preeclampsia and preterm birth, in which VEGF has established roles^35^. This cohort was also focused on nulliparous women and had less genetic ancestry diversity. Detailed characterization of this locus can provide further insight into how it regulates VEGF levels throughout pregnancy, both in terms of temporal changes and connections to parity, genetic ancestry, or pregnancy outcome.

Another advantage of measuring two timepoints is the opportunity to study changes in levels of these biomarkers and the extent to which genetics contributes to modulating these changes. For Δ, we identified 7 biomarkers with significant heritability highlighting a role for genetic variation in the dynamics of immune response over pregnancy, perhaps analogous to the unique genetics of response to immune challenge^17–19^. We also identified GWS associations for 9 biomarkers’ Δ. These loci are non-overlapping with T1/T2 associations and enriched in distinct biological processes, primarily developmental, in comparison to immune signaling enrichment in T1/T2. Another trend we observed with Δ-associated loci was the enrichment of transcription factors, several of which have already been shown to regulate expression of various immune biomarkers^36^. For instance, *IL12B* is a known target of transcription factor *RXRA*, which has a suggestive association with IL-12p70^36,37^. Additionally, *RXRA* and other transcription factors such as *MAF* and *RORA* have known roles in progression of pregnancy and pregnancy complications^27,38,39^. These could imply transcriptional mechanisms guiding the dynamics of pregnancy immune changes.

Our previous study in the EMA cohort is the only other available study of maternal genetics and immune biomarkers during pregnancy. Comparison with summary statistics showed limited overlap, which could be driven by differences between studies. The EMA cohort was smaller (n=790), which limits its power in comparison to this study, and sampled from a southern (vs. northern) California population. It examined a subset of maternal biomarkers at only one timepoint (similar to T2). Ultimately, 6 of 20 biomarkers compared had at least one suggestive SNP also nominally significant in the current study, representing ∼6% of independent loci at *P*<0.05. The EMA study also included fetal genetics and biomarkers, revealing a strong influence of fetal genetics on maternal biomarkers and *vice versa*. One neonate locus near *PLCL2* was associated with neonatal levels of MIG and 10 other biomarkers. In this current study, we also identified a MIG-associated suggestive SNP mapped to *PLCL2*. This could indicate overlap between maternal and fetal genetic regulation. A limitation of our current study is that we have neither fetal genetics nor neonatal biomarker measurements to ensure specificity of maternal effects.

Throughout pregnancy, the placenta and its surrounding environment undergo transformations to support placentation and fetal development, including changes in the milieu of maternal immune cells in the uterus. A study on spatiotemporal dynamics of maternal-fetal interface^40^ characterized a strong relationship between gestational age and maternal immune cell composition. They predominantly observed natural killer cells and T cells with immunosuppressive phenotypes at 6-8 weeks. Tolerogenic macrophages and extravillous trophoblasts became the dominant cell types by weeks 16-20. Likewise, in our data the expression signatures of natural killer cells and T cells were enriched for T1 associations in contrast to macrophages and T cells at T2.

The maternal immune environment and its dysregulation have been linked to adverse pregnancy conditions, although the extent to which genetic factors causally contribute to this relationship is not understood^41,42^. Our investigation of overlap between genetics of immune mediators and pregnancy complications revealed 20 biomarkers enriched for PE, PTB, and GDM-associated loci. Some of the biomarkers our genetic data identified have already been studied in these conditions, such as the relationship of IFNγ and IL-10 with preeclampsia^43,44^ and the possibility of IP-10 connecting inflammatory anti-angiogenic states^45^, all three of which showed an enrichment of PE loci. Mendelian randomization studies support a causal link implied by our data between IL-7 and IL-12p70 levels and GDM^46,47^. We observed enrichment of PE, PTB, and GDM-associated loci in IL-4, and disruption of its anti-inflammatory role in pregnancy has been linked to adverse outcomes^48^. G-CSF has roles in embryo implantation^49^ and is being explored as a therapeutic option for recurrent pregnancy loss^50^. It showed an enrichment of PE and PTB loci, along with associations with known pregnancy complication genes *CSF3* and *HLA-DQA1*, suggesting that genetic autoregulation could be critical in maintaining healthy levels of G-CSF. However, it’s worth noting that there were also biomarkers with critical roles in maternal immune environment, such as IL-6^51^ and IL-17A^52^, where we didn’t observe this genetic overlap or any GWS signals, potentially implicating non-genetic factors more strongly in these associations. Follow-up investigation could help positively confirm ‘environmental’ contributions for the remaining inflammatory markers and nominated pregnancy outcomes. Disruption of inflammatory protein levels could also affect the health and neurodevelopmental outcomes of the baby^53,54^. Therefore, future studies may be able to use our genetic results to disentangle causal relationships between immune dysregulation, pregnancy complications, and neurodevelopmental outcomes.

To remain unbiased and consistent while mapping SNPs to nearby genes, we used annotation methodology prioritizing distance to the nearest transcription start site. However, there may be better candidate genes in the same region. For instance, fractalkine-associated rs1051112 was assigned to *TMEM121*, but this SNP is also near *IGHG3* which encodes an immunoglobulin heavy chain. IL-15-associated rs112022139 was mapped to *SUV39H2,* but nearby *DCLRE1C* has roles in V(D)J recombination. TGFα-associated rs10918270 was annotated to *OLFML2B*, though it is in the intronic region of *ATF6*, an important arm of unfolded protein response and ER stress which contributes to preeclampsia^55^. rs73043012, which is a pregnancy unique variant associated with T2 only, was assigned to *CATSPERG*, involved in sperm cell hyperactivation. Regional eQTLs act on *CATSPERG* and *SPINT2*^56^. *SPINT2* is an inhibitor of HGF activator, which is highly expressed in endometrial epithelial cells (enriched in T2-implicated genes). Thorough finemapping across GWS loci may lead to additional insights into determination of the immune repertoire in pregnancy.

Our study has limited power for detecting heritability and GWS signals. Certain confounders may further limit power, including subclinical infections or immune disorders. The lack of pre-pregnancy or post-partum samples limited our ability to assess which variants represent pregnancy-specialized associations. As an alternative, we focused on comparisons with existing studies in the general non-pregnant population. However, because our cohort is female-only, some of the observed differences could be driven by sex (or other) differences between cohorts and not specific to pregnancy. Despite these (and other) limitations, our observations provide an examination of variants associated with immune biomarkers in early and mid-pregnancy along with genetic drivers of change observed over pregnancy, nominate putative pregnancy-unique associations, and suggest a relationship between genetics of maternal immune millieu and adverse pregnancy outcomes.

## METHODS

### Study population

The detailed description of the IMPaCT (Immune and Metabolic Markers during Pregnancy and Child Development) cohort is available at Croen *et al*.^20^. Briefly, participants were selected from children born at Kaiser Permanente Northern California (KPNC) from January 2011 to January 2016 and enrolled in the Research Program on Genes, Environment, and Health (RPGEH) pregnancy cohort^57^, including donation of a blood sample during the first and second trimesters of pregnancy and permission to access their own and their child’s KPNC electronic health records (EHRs) for future studies. In December 2019, pregnancies resulting in cases of ASD (n = 339), DD (n = 1193), and GP controls (n = 904) were selected for study. Study procedures were approved by the KPNC Institutional Review Board. The self-reported maternal race distribution of the study participants was as follows: 45.7% white, 24.6% hispanic, 20.7% asian, 5.4% black, and 3.6% other or unknown. Population structure was further explored by computing genetic principal components (PCs) using PLINK^58^ [Supp Fig 5a].

### Genotyping and immune biomarker measurements

Maternal blood samples were collected during the first and second trimesters of pregnancy. The detailed description of sample preparation and genotyping procedures is available at Croen *et al.*^20^. In brief, genotyping was performed on the Axiom Precision Medicine Research Array (PMRA) by the Genomics Core Facility at UCSF, with a standard quality control pipeline. Out of approximately 800,000 markers that were genotyped, 727,655 markers passed quality control. Imputation was performed using the Michigan Imputation Server^59^ with the 1000 Genomes Reference panel^60^ resulting in 3,0969,804 markers, with additional quality controls in place.

Concentrations of the following serum and plasma biomarkers were measured using Milliplex human cytokine/chemokine/growth factor 48-plex bead kit (HCYTA-60K-PX48) following the manufacturer’s instructions: sCD40L, epidermal growth factor (EGF), eotaxin, fibroblast growth factor 2 (FGF-2), Fms-related tyrosine kinase 3 ligand (FLT-3L), fractalkine, granulocyte colony-stimulation factor (G-CSF), granulocyte macrophage colony-stimulating factor (GM-CSF), GRO1 oncogene (GROα; CXCL1), interferon alpha 2 (IFNα2), IFNγ, interleukin (IL)-1α, IL-1β, interleukin-1 receptor antagonist (IL-1RA), IL-2, IL-3, IL-4, IL-5, IL-6, IL-7, IL-8, IL-9, IL-10, IL-12p40, IL-12p70, IL-13, IL-15, IL-17A, IL-17E/IL-25, IL-17F, IL-18, IL-22, IL-27, interferon gamma-induced protein 10 (IP-10), monocyte chemoattractant protein-1 (MCP-1), MCP-3, macrophage colony-stimulating factor (M−CSF), macrophage-derived chemokine (MDC; CCL22), monokine induced by gamma (MIG; CXCL9), macrophage inflammatory protein-1 alpha (MIP-1α), MIP-1β, platelet-Derived Growth Factor AA (PDGF AA), PDGF AB/BB, regulated upon activation, normal T cell expressed and secreted (RANTES; CCL5), transforming growth factor alpha (TGFα), tumor Necrosis Factor alpha (TNFα), TNFβ, and vascular endothelial growth factor (VEGF).

### Data processing

Data processing and analysis steps described below were performed in R (v4.4.1) unless specified otherwise. There were 2227 individuals with first (T1) and 2098 individuals with second (T2) measurements available, as we only had one measurement available for a subset of individuals. Values below the limit of detection were treated as missing values. The median of percentage of missing values across biomarkers was 1.1% for T1 and T2. However, 11 biomarkers at T1 and 10 at T2 had more than 10% missing values. This did not lead to a non-normal distribution of these biomarkers’ measurements, nor genomic inflation in QQ plots. IL-3 measurements were excluded from further studies due to a high rate of missing values below the limit of detection (∼85%).

Each biomarker’s T1 and T2 measurements were log transformed to normalize the distributions. Gestational age in days when samples were collected varied across individuals and covered a wide range within first and second trimesters [Supp Fig 5b]. On average, gestational age at first sample collection was 62.9 (sd: 15) and second sample collection was 121.8 (sd: 19). To account for variation associated with this, linear regression of gestational age against log-normalized T1 and T2 measurements was used to obtain residuals. These gestational age corrected values were then used as the final T1 and T2 values in further analyses. However, IL-27 and VEGF T1 measurements displayed a non-linear trend across gestational age. Therefore, we used piecewise linear regression (segmented R package v2.1.4) to better account for their temporal trends. Finally, for each biomarker, values +/− 3 standard deviation away from mean were excluded as outliers. The average number of outliers excluded was 24 for T1 and 22 for T2, and the maximum number of outliers excluded was 49 for T1 and 41 for T2.

To create a variable that reflects the changes between two measurements (Δ), we computed the difference between gestational age corrected T1 and T2 measurements. This variable was computed for the subset of individuals with two measurements that were obtained at least two weeks apart from each other, corresponding to 1892 individuals. The average gap between two measurements was 59 days (sd: 21.2). Values +/− 3 standard deviation away from mean were excluded as outliers. The average number of outliers excluded was 29 and the maximum number of outliers excluded was 67.

### SNP-based heritability

We used Restricted Maximum Likelihood (REML) model implemented in GCTA software^61^ to estimate SNP-based heritability for each biomarker and time variable, accounting for the following covariates: maternal education, maternal age, maternal race, fetal sex, birth year, child outcome, measurement plate number, and the first ten PCs. While comparing results across time variables, *h^2^*>0.5 was defined as high heritability estimate and 0.2<*h^2^*<0.5 was defined as moderate.

### Genome-wide association study

We performed GWAS using PLINK software^58^ per biomarker and time variable combination, accounting for the following covariates: maternal education, maternal age, maternal race, fetal sex, birth year, child outcome, measurement plate number, and the first ten PCs. With the Δ variable, the difference between gestational age at second and first measurements was also included as a covariate to account for the variation in gap between measurements. SNPs with minor allele frequency less than 1% were excluded. QQ plots generated for quality control were in line with expected distributions. METAL^62^ was used to test whether T1 and T2 effect size estimates of GWS SNPs were significantly different.

We used 5×10^−8^ as the p-value threshold for GWS SNPs and 10^−6^ as suggestive threshold, unless specified otherwise. GREAT online portal^63^ was used to annotate SNPs to nearby genes using “single nearest gene” option with default parameters (the extension limit of 1000kb). Variants that were not annotated to any gene with this method were not included in gene-based analyses.

### GWAS permutations

To create a set of permuted GWAS runs, we generated randomized matrices of biomarker levels. We first created a combined matrix of T1, T2, and Δ values along with all covariates except for 10 PCs by concatenating each individual’s corresponding values. Individual IDs were then shuffled to create a randomized matrix. This preserves any relationship between biomarkers within an individual by keeping all measurements obtained from one individual together. 100 different randomized matrices were generated, and they were then separated into T1, T2, and Δ files to perform GWAS with each permuted dataset following the same protocol described above.

Suggestive SNPs within a given biomarker’s permuted run were obtained by selecting the same number of SNPs that passed the suggestive threshold in the original result, after sorting the permuted results based on p-values. This was done separately for each individual permuted result, generating 100 lists of permuted suggestive SNPs per biomarker.

### Pathway enrichment analysis

After mapping suggestive SNPs (*P*<10^−6^) to genes, we obtained the union of genes associated with all biomarkers to create a collective set of genes for T1, T2, and Δ separately. These were used to run pathway enrichment analysis with clusterProfiler^64^ (v4.12.0). MSigDB^65^ was used to obtain gene sets of GO-Biological Process and GO-Molecular Function (v2025.1). We also mapped every SNP that was included in GWAS to genes in order to generate a background gene set (17612 genes) for enrichment. TFlink database^36^ was used to obtain the list of transcription factors, based on annotations obtained from small-scale experiments.

### Comparison against existing GWAS on immune biomarkers

Full summary statistics of study 1 by Ahola-Olli *et al*.^14^ (https://www.ebi.ac.uk/gwas/publications/27989323) and study 2 by Zhao *et al*.^15^ (https://www.ebi.ac.uk/gwas/publications/37563310) were downloaded from GWAS catalog.

For this analysis, instead of using the suggestive threshold of 10^−6^, we tested a range of p-value thresholds to identify a value that would maximize the coverage of shared associations and limit the number of possible false positive unique associations. 10^−7^ provided an optimal solution, as relaxing the threshold beyond this value did not lead to the inclusion of any additional shared associations. Therefore, this threshold was used to identify the list of highly suggestive SNPs in both our study and comparison studies.

The following steps were performed per biomarker with a corresponding summary statistic available in study 1, separately for T1 and T2. We first clumped highly suggestive SNPs (*P*<10^−7^) using PLINK^58^ (with default parameters, except for clump-r2=0.2 and clump-kb=500). We then identified the subset of clumps that is also present in study 1. If the index SNP itself was not present, we checked the remaining SNPs that are part of the clump and if at least one SNP was found in the other study, the given clump was counted as an overlapping clump. Afterwards, we looked into the p-value of these clumps in study 1 to divide them into the “shared” or “unique” category based on the same 10^−7^ threshold. This process was then repeated for study 2.

The Manhattan plots include all highly suggestive SNPs that are present in both our and comparison study and additionally highlight the corresponding index SNPs of each clump. Genes that were assigned to each region are also annotated along with the biomarkers they are associated with, matching the color code of corresponding chromosomes. These plots were generated using ggmanh R package (v1.8.0).

An additional comparison was performed against our previous EMA study with a pregnancy specific cohort^16^. These were done only with T2 results as EMA cohort’s sample collection time frame was within the second trimester. We focused on 20 of the biomarkers that were available in the maternal EMA dataset. We then identified the list of independent loci below a suggestive threshold (*P*<10^−5^) in EMA that is also nominally significant (*P*<0.05) in T2 (6 of 99).

### Comparison against placental gene expression

Suryawanshi *et al.*^25^ performed scRNA-seq on first trimester placenta, identifying 13 maternal and 9 fetal cell types; we focused on the maternal cells. For each cell type, we first ranked genes based on their expression level (with highest expression getting the first rank) to make results across cell types more comparable. Per time variable, we then identified the rank distribution of genes assigned as nearest to suggestive SNPs and determined the rank that corresponds to the 10^th^ percentile of this distribution. This rank was used as a threshold to compare against each permuted GWAS result. We computed the number of times where a permutation’s percentage of genes with ranks lower than this threshold was greater than 10% to estimate a permutation p-value.

We downloaded “RNA expression levels per gene and cell type” dataset from Human Protein Atlas^26^ website and repeated the analysis described above with the cell types that most closely resemble the origin of placental cell types.

### Comparison against pregnancy complication studies

Barbitoff *et al.*^27^ curated genes associated with various pregnancy complications based on multiple sources. We specifically focused on the Public Health Genomics and Precision Health Knowledge Base (PHGKB v6.2.1) HuGE Navigator curation. Using the union of annotated genes per time variable, we identified the number of genes known to be associated with PA, PE, GDM, and PTB within the full set of curated genes.

To perform comparisons against GWAS related to these complications, we obtained full summary statistics of studies focusing on PE by Tyrmi *et al.*^28^ (https://www.ebi.ac.uk/gwas/publications/37285119), GDM by Elliott *et al.*^29^ (https://www.ebi.ac.uk/gwas/publications/38182742), and PTB by Sole-Navais *et al.*^30^ (egg-consortium.org/Gestational-duration-2023.html). The following steps were performed against each of these studies per biomarker and time variable combination. We first identified suggestive SNPs (*P*<5×10^−5^) in the comparison study that is also available in our study and clumped these SNPs. Afterwards, we computed the percentage of these independent loci that also has nominally significant (*P*<0.05) associations with a given biomarker. This step is then repeated with 100 permuted GWAS results to estimate significance for the level of overlap observed.

## Data Availability

All data produced in the present study are available upon reasonable request to the authors.

## ACKNOWLEDGEMENTS

This work was supported by the National Institute of Child Health and Human Development (Grant No. R01HD095128 and R01HD112326). The findings and conclusions in this report are those of the authors and do not necessarily represent the official position of the National Institutes of Health.

Data used in this study were provided by the Kaiser Permanente Research Bank from the Kaiser Permanente Research Bank collection, which includes the Kaiser Permanente Research Program on Genes, Environment, and Health, funded by the Robert Wood Johnson Foundation, the Wayne and Gladys Valley Foundation, The Ellison Medical Foundation, and the Kaiser Permanente Community Benefits Program. Access to data used in this study may be obtained by application to the Kaiser Permanente Research Bank at kp.org/researchbank/researchers.

We thank Susan Fisher and Andrea Sandoval for their helpful comments and contributions. The authors report no biomedical financial interests or potential conflicts of interest.

## AUTHOR CONTRIBUTIONS

LAW, JVW, and LAC conceived the project. LAC designed and identified the pregnancy cohort and sampling. JVW provided measurements of immune biomarkers. MT and LAW oversaw genotyping, QC, and preparation of the genetic dataset. MC and LAW designed the formal analysis steps. MC performed the data analysis. MC and LAW wrote the original draft. All authors contributed to review and editing and approved the final manuscript for publication.

---

**Supp Table 1.** Δ-associated SNPs (*P*<10^−6^) annotated to transcription factors. If there are multiple SNPs annotated to the same gene, only the most significant SNP is displayed.

| Biomarker | CHR | Gene (TF) | SNP | $\Delta P$ |
| --- | --- | --- | --- | --- |
| IL-1β | 7q21.3 | *BHLHA15* | rs73151168 | $6.16 \times 10^{-7}$ |
| GM-CSF | 2p21 | *EPAS1* | rs79824099 | $8.07 \times 10^{-7}$ |
| IL-13 | 7p22.1 | *FOXK1* | rs78685992 | $7.93 \times 10^{-7}$ |
| TGFα | 7p22.2 | *FOXK1* | rs28609201 | $2.99 \times 10^{-7}$ |
| IL-1α | 6p25.3 | *FOXQ1* | rs114247784 | $1.73 \times 10^{-7}$ |
| IL-15 | 3q29 | *HES1* | rs4686663 | $1.92 \times 10^{-7}$ |
| GM-CSF | 16q23.1 | *MAF* | rs12444900 | $4.89 \times 10^{-7}$ |
| TNFβ | 5q14.3 | *MEF2C* | rs145965369 | $4.76 \times 10^{-7}$ |
| Eotaxin | 8q13.1 | *MYBL1* | rs547031641 | $8.46 \times 10^{-7}$ |
| IP-10 | 14q13.3 | *PAX9* | rs4904582 | $1.48 \times 10^{-8}$ |
| MIP-1α | 15q22.2 | *RORA* | rs1370432 | $1.49 \times 10^{-7}$ |
| IFNγ | 6p21.1 | *RUNX2* | rs78548485 | $1.74 \times 10^{-7}$ |
| IL-12p70 | 9q34.2 | *RXRA* | rs3132301 | $2.65 \times 10^{-7}$ |
| M-CSF | 18q23 | *SALL3* | rs34653042 | $7.63 \times 10^{-7}$ |
| RANTES | 18q23 | *SALL3* | rs878916908 | $5.01 \times 10^{-7}$ |
| M-CSF | 2p25.2 | *SOX11* | rs73913776 | $8.67 \times 10^{-7}$ |
| IL-12p40 | 2p25.2 | *SOX11* | rs1389491716 | $3.25 \times 10^{-7}$ |
| FGF-2 | 12p12.1 | *SOX5* | rs544998569 | $4.16 \times 10^{-7}$ |
| FLT-3L | 22q13.31 | *TBC1D22A* | rs16996074 | $5.34 \times 10^{-7}$ |
| TNFα | 15q23 | *TLE3* | rs11072135 | $8.63 \times 10^{-7}$ |
| IL-1α | 10q24.31 | *TLX1* | rs10128244 | $5.11 \times 10^{-8}$ |
| IL-2 | 19q12 | *URI1* | rs75515131 | $1.65 \times 10^{-7}$ |
| IL-6 | 17q22 | *VEZF1* | rs62081792 | $3.06 \times 10^{-7}$ |
| IL-1α | 2q22.3 | *ZEB2* | rs74468904 | $6.17 \times 10^{-7}$ |

**Supp Fig 1.**
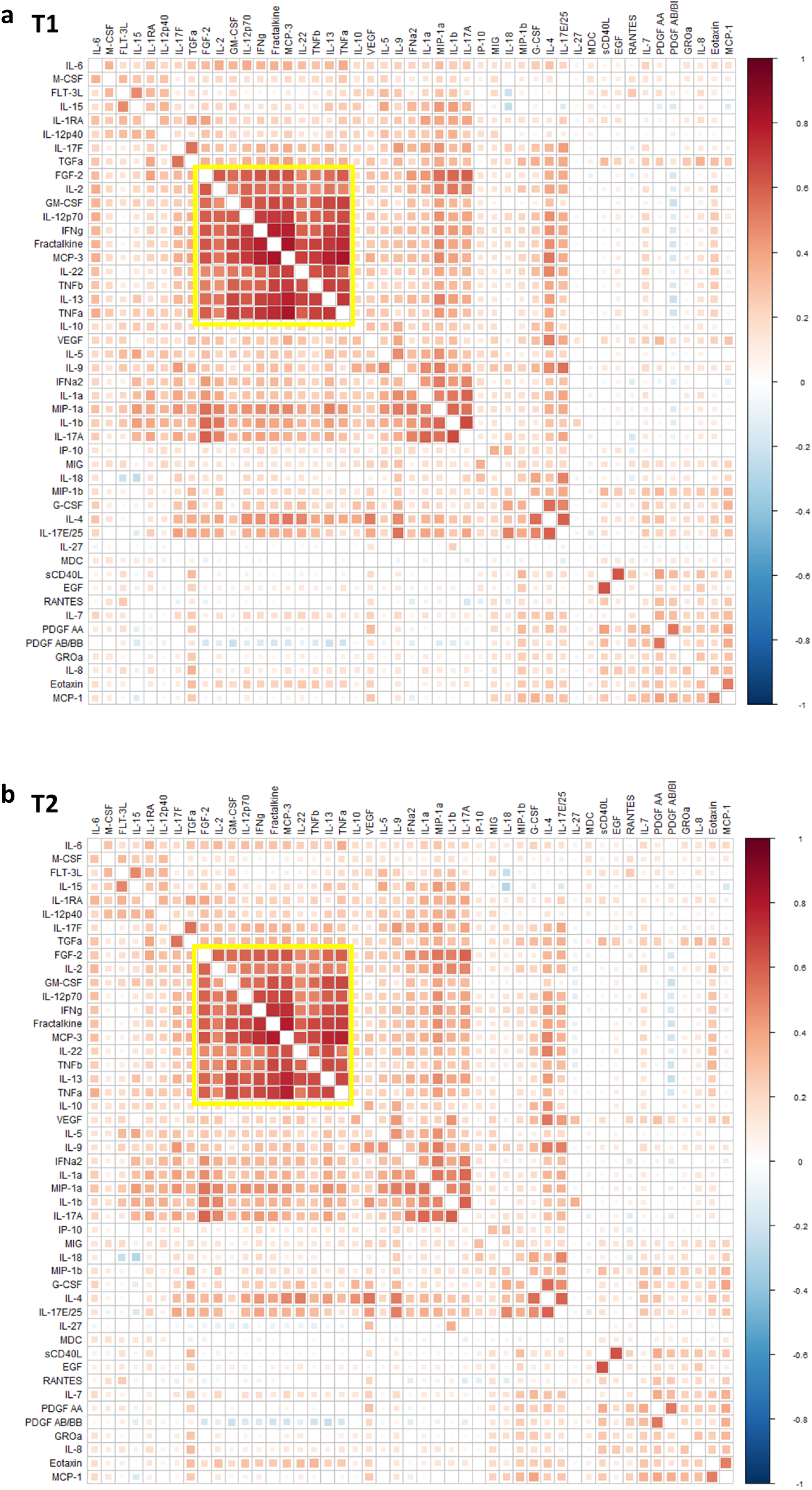
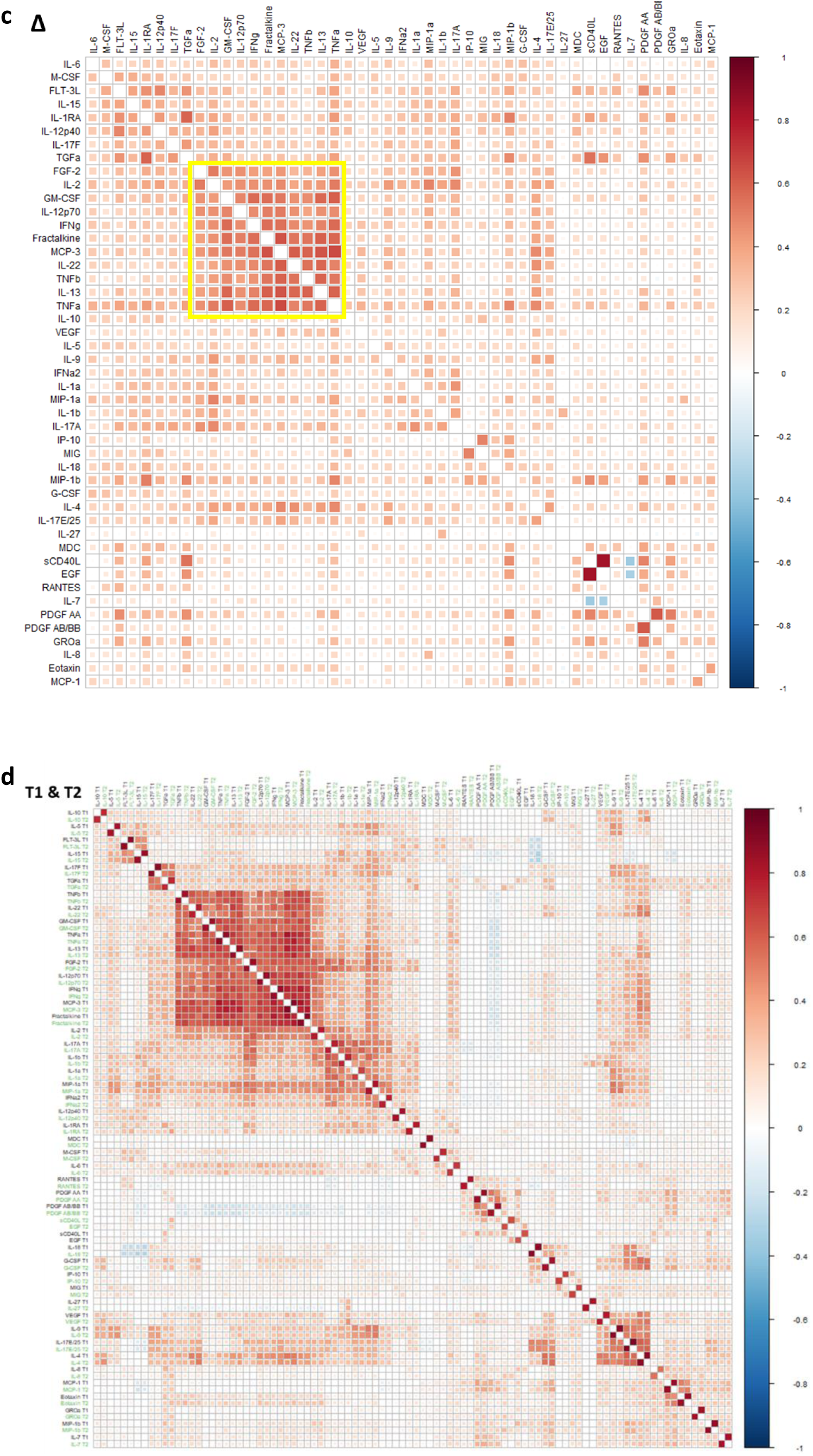
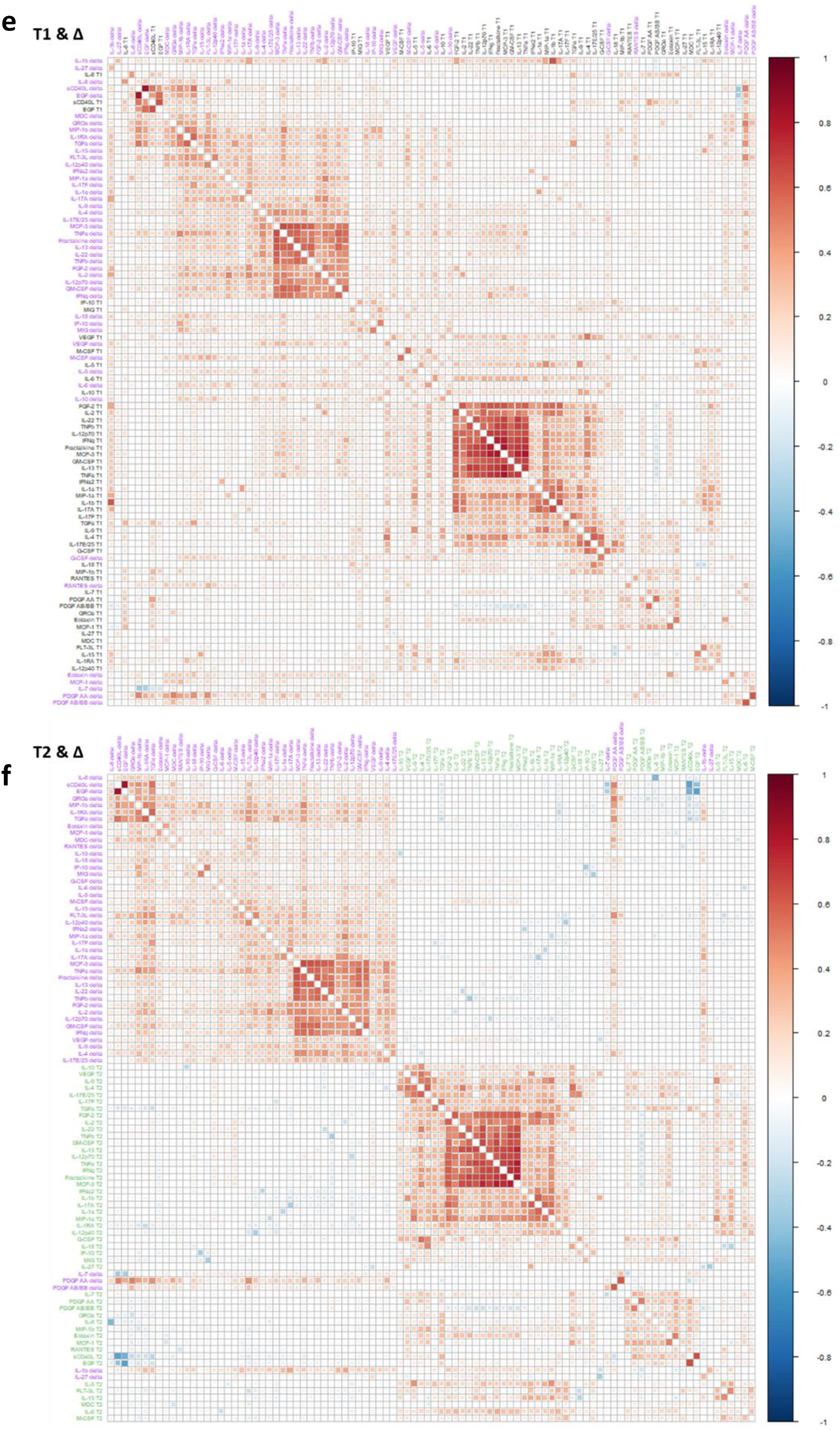
Biomarker correlation. **a-c**. Pairwise correlation matrices of T1 (a), T2 (b), and Δ (c). T1 heatmap (red – positive correlation; blue – negative correlation) is ordered based on hierarchical clustering, and biomarkers in T2 and Δ heatmaps are presented in the same order for comparison. The yellow square highlights 11 highly correlated biomarkers. **d-f.** Pairwise correlation matrices of T1 & T2 (d), T1 & Δ (e), and T2 & Δ (f). T1 labels are shown in black, T2 in green, and Δ in purple.

**Supp Fig 2.**
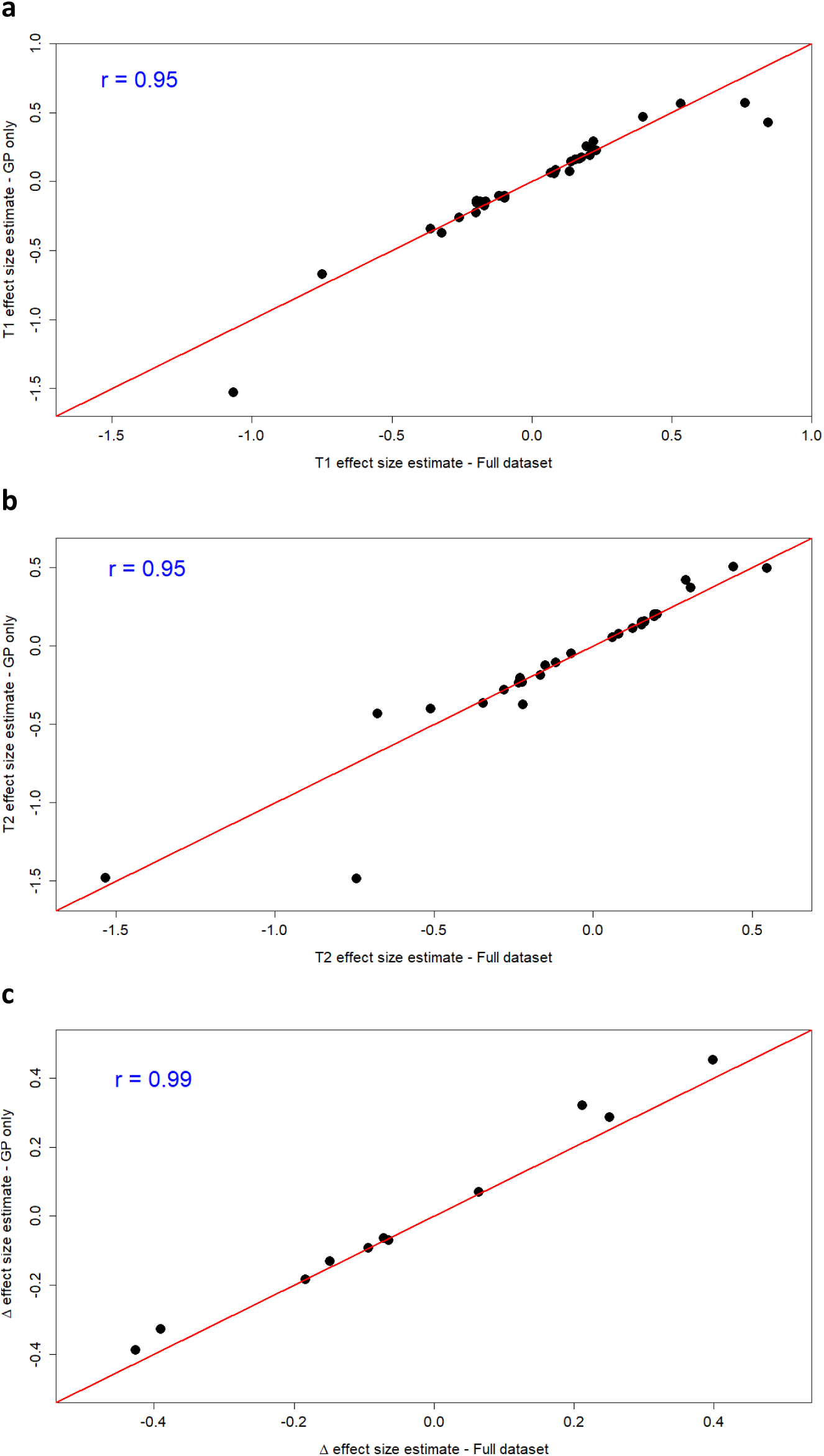
Neurodevelopmental Outcome Sensitivity Analysis. Effect size estimates of T1 (a), T2 (b), and Δ (c) GWS SNPs are plotted against estimates obtained when GWAS is repeated with GP-only samples, excluding pregnancies with ASD or DD outcomes. Correlation coefficients are displayed on the plots.

**Supp Fig 3.**
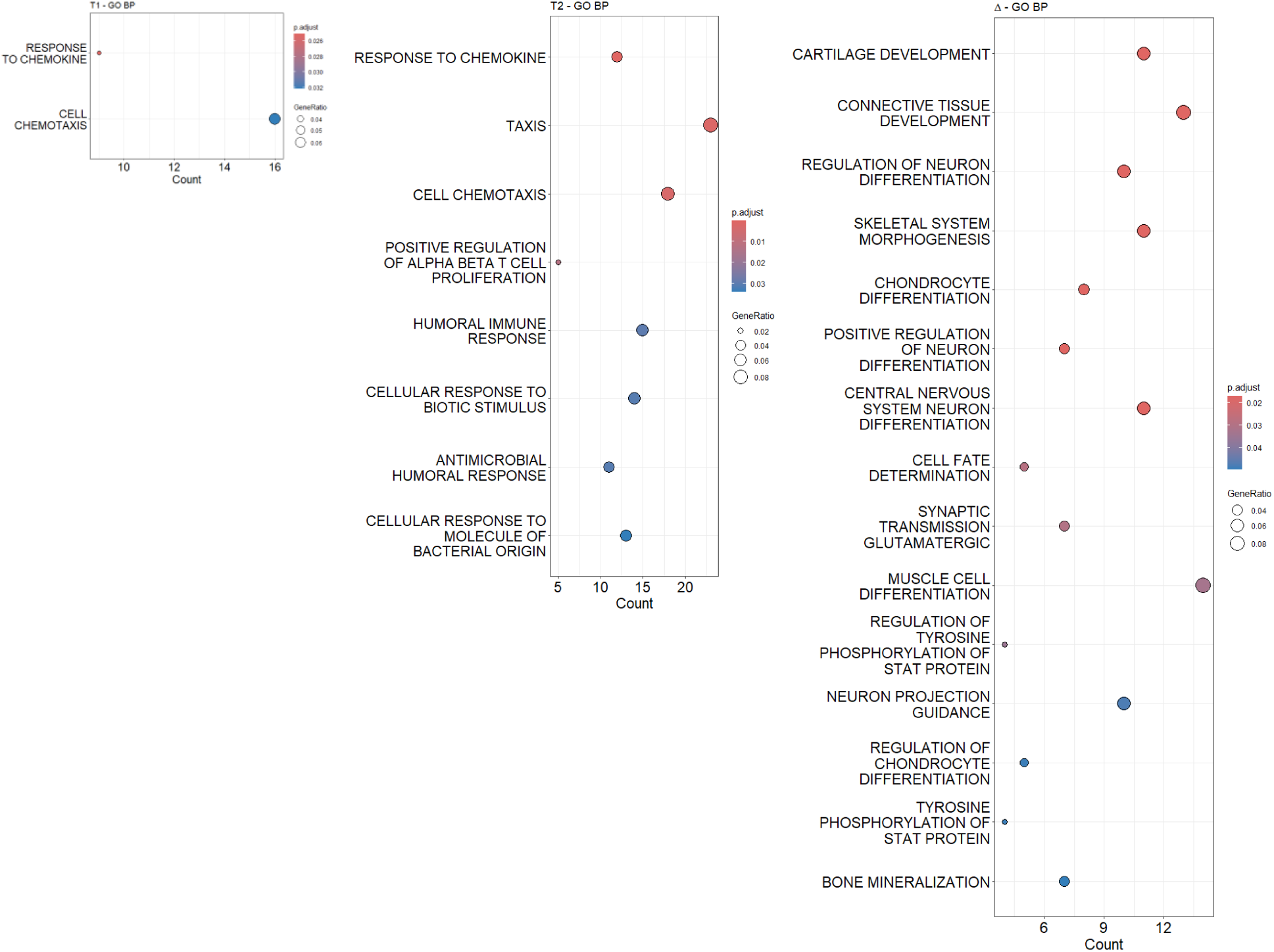
Gene Ontology Enrichment. Significantly enriched GO-BP pathways within each time variable’s set of query genes assigned to nearby suggestive SNPs (*P*<10^−6^). Categories are ordered based on adjusted p-values and circles are color coded based on adjusted p-values. Count in x axis denotes the number of genes in the query set that belong to the category. Circle sizes are proportional to gene ratio, which denotes the ratio of the number of genes in the query set in that category to the total number of genes in the category.

**Supp Fig 4.**
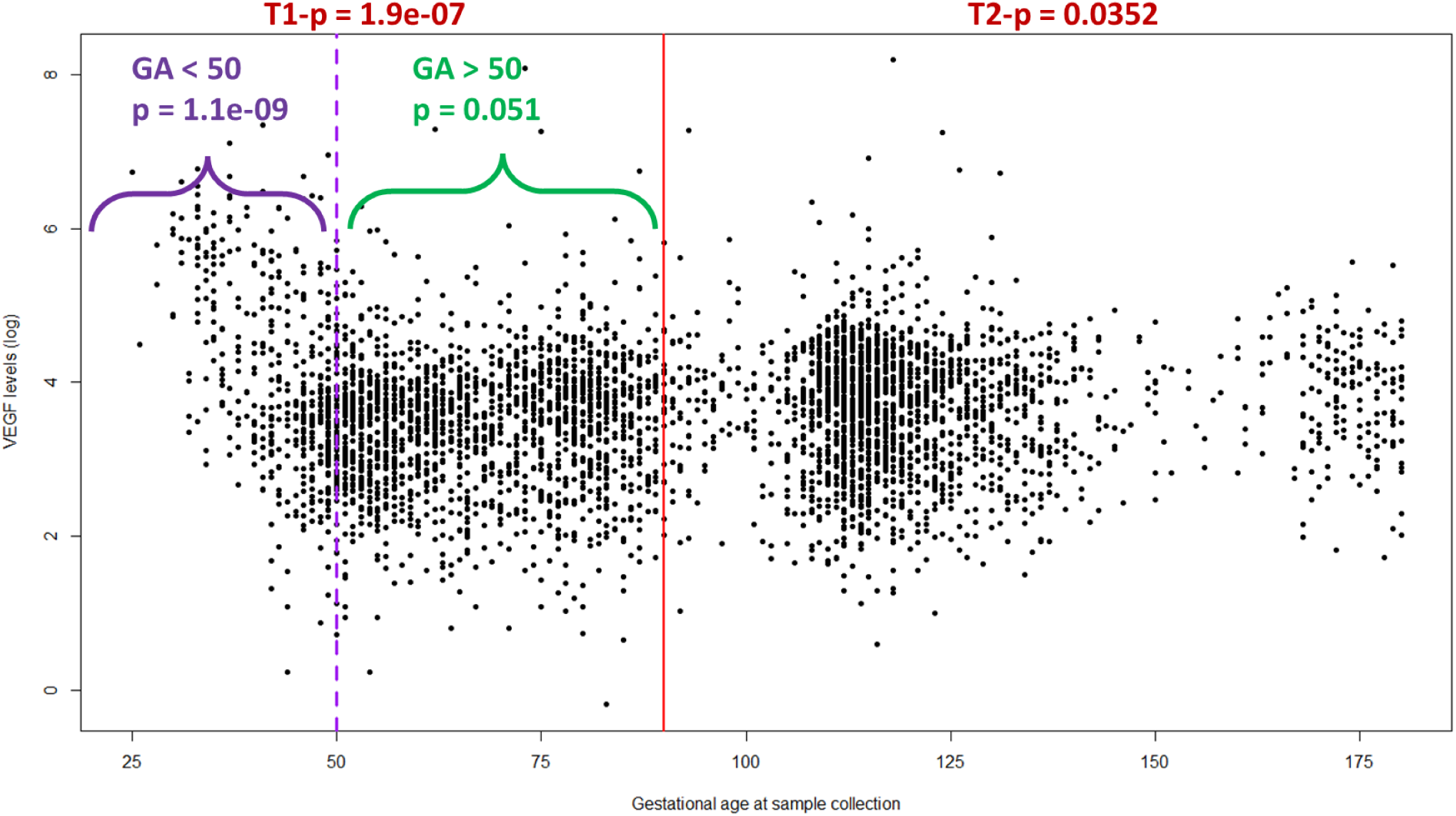
VEGF-rs6921438 Association by Gestational Age. Log-normalized VEGF level from each sample is plotted against the corresponding gestational age. Red line indicates 90 days, the threshold between first and second trimesters. Dashed purple line indicates 50 days. T1 data were divided based on this 50-day timepoint and p-values of the association between rs6921438 and VEGF levels within each group are shown in purple and green. p-values of the association between rs6921438 and VEGF levels within full T1 and T2 datasets are shown in red.

**Supp Fig 5.**
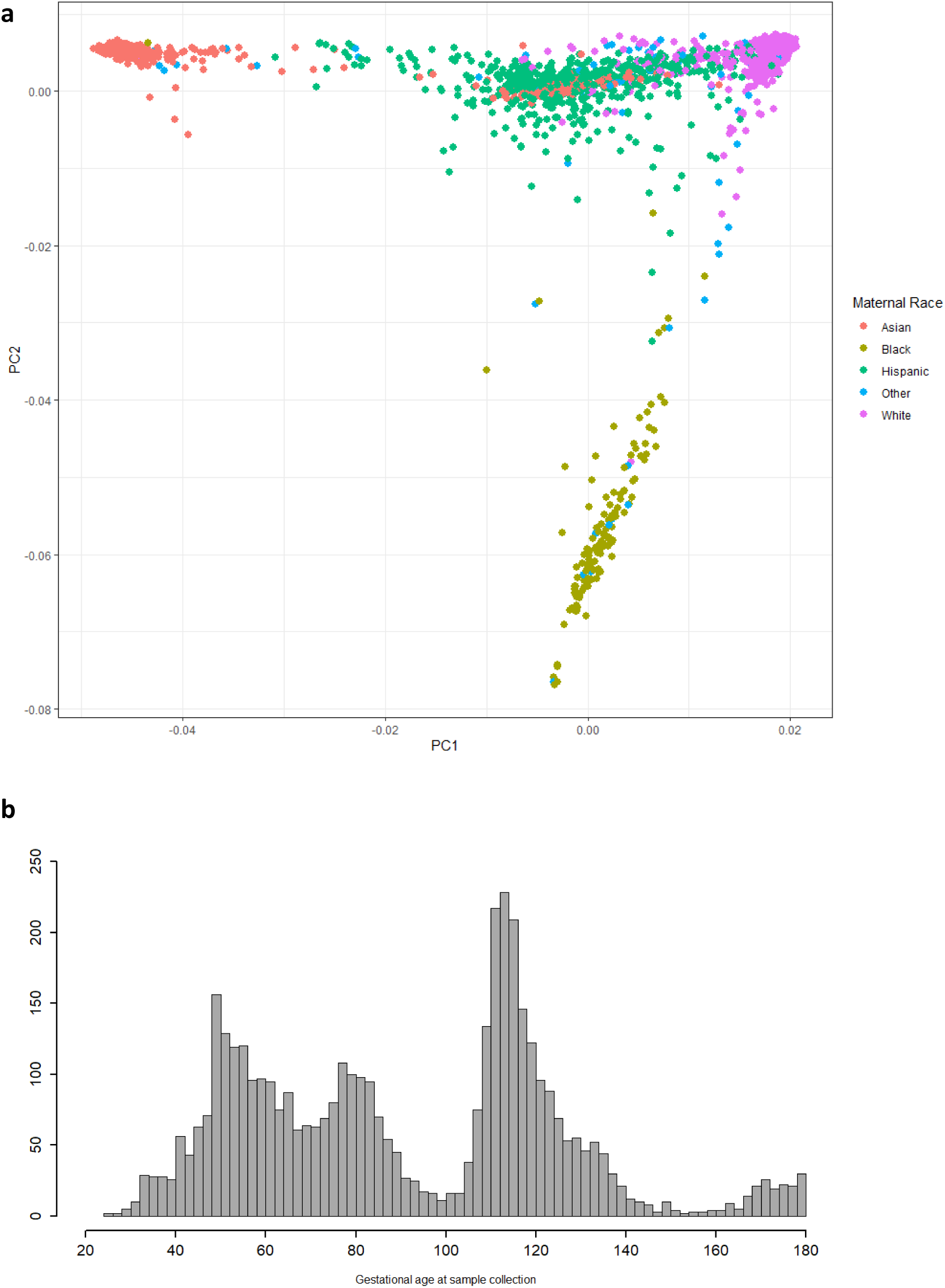
Data Description. **a.** Genetic principal components PC1 and PC2 are plotted to visualize genetic ancestry. Dots are colored based on self-reported maternal race/ethnicity. **b.** Histogram of gestational age in days at sample collection.

